# Effective tumor kinetics inferred from single routine H&E biopsies enable counterfactual virtual radiotherapy trials

**DOI:** 10.64898/2026.09.10.26362768

**Authors:** Pirmin Schlicke, Caner Ercan, Sara Ranjbar, Ashlee N. Seldomridge, Sadhna Aggarwal, Adwait Mehta, Tania P. Sainz, Mohammad U. Zahid, Penny Fang, Veronika A. Hofmann, Stefano Pasetto, Tatiana Cisneros Napravnik, Nahum Puebla-Osorio, Christina Kuttler, Ann Klopp, Lauren Colbert, Ashley M. Holder, Roi Weiser, Steven H. Lin, Carlos A. Torres-Cabala, Francisco Vega, Yinyin Yuan, Heiko Enderling

## Abstract

Routine H&E-histopathology captures spatial snapshots of tumor ecosystems, yet computational pathology largely treats them as textures without explicit links to underlying kinetics. We show that patient-specific parameters of a mechanistic reaction-diffusion model can be inferred from a single biopsy. Cell-nucleus point patterns are fitted jointly to the analytical two-point correlation function and power spectral density, recovering patient-specific mechanistic proliferation and diffusion rates across eleven multicenter cohorts. Derived dispersion length and front velocity define mechanistic phenotypes stratifying progression-free and overall survival and adding information independent of routine covariates. We illustrate counterfactual radiotherapy simulations in glioblastoma and show that a virtual clinical trial deescalating histology-calibrated faster-growing tumors to hypofractionation while dose-intensifying slower-growing tumors adds 96.7 days of restricted mean progression-free *in silico* survival over standard of care (permutation *p* < 0.001 against random allocation to the same arms). This provides a low-barrier transition from routine histopathology to mechanistically interpretable model-based forward simulations.

## Introduction

Routine histopathology is a standard preclinical and clinical methodology for disease diagnosis that characterizes the spatial architecture of a tumor and its microenvironment, thereby providing a snapshot in time of a complex dynamic system. Recent advances in digital pathology, especially the implementation of artificial intelligence, have provided novel clinically actionable insights that can improve cancer diagnosis, prognosis, and treatment. Deep learning models trained on hematoxylin and eosin (H&E)-stained slides and slide labels have achieved impressive clinical performance in diagnosis and the prediction of genomic alterations, survival, and treatment outcomes^1–6^. Recently, multimodal model architectures that integrate histology with proteomic and genomic profiles in pan-cancer cohorts have been shown to jointly capture molecular status, morphology, and outcome^7–11^. However, most of these methods treat histologic features as high-dimensional texture and learn features specific to the designed task, while the explicit relationship between these features and the underlying tumor kinetics and tissue-scale dynamics remains obscure.

Mechanistic mathematical models have demonstrated that the growth and spatial spread of solid tumors can often be formalized using low-dimensional dynamical systems. For example, in diffuse glioma, reaction-diffusion equations calibrated from longitudinal magnetic resonance imaging (MRI) images have been used to estimate patient-specific tumor proliferation and diffusion rates to predict tumor growth and recurrence and to rationalize the effect of radiotherapy and surgery on survival^12–18^. These studies show that a small number of kinetic parameters can summarize complex tumor dynamics across spatial and temporal dimensions and may serve as basis for imaging-driven digital tumor twins^19–22^. The generalizability of these advances is limited, however. MRI-based calibration is restricted to disease sites with standardized volumetric imaging and does not naturally extend to many cancers where pre-treatment histopathology is the primary diagnostic readout.

Spatial statistics provide a complementary approach to MRI-based calibration by exploring tissue organization at cellular resolution. The pair correlation function and the related two-point correlation function (2PCF) have long been used in physics, astronomy, and other scientific fields to quantify dispersion and clustering in point processes^23–27^. Recent works have adapted these tools to digital pathology, introducing spatial point patterns of individual cells to characterize tumor-to-immune cell organization and to derive spatial biomarkers associated with patient outcomes^28–30^. By design, these analyses are descriptive: they quantify spatial structures but are not linked to explicit dynamical models that can be calibrated to underlying dynamics and employed for forward simulations. Complementing this line of work, Somer et al. recently demonstrated that dynamical models of cell-population change can be inferred from a single spatial proteomics sample through one-shot tissue dynamics reconstruction^31^. Their approach derives division-rate dependencies on cellular neighborhood composition and constructs a data-driven dynamical model of cell populations. A conceptually related question was addressed from a complementary angle, whether the kinetic parameters of an explicit, closed-form reaction-diffusion partial differential equation can be inferred directly from routine H&E-stained tissue, the single diagnostic modality available for nearly every solid-tumor patient.

In this work, we introduce histopathology-derived tumor dynamics (HDTD), a framework that directly calibrates the mechanistic reaction-diffusion equation, and thereby patient-specific cancer dynamics, from a single, routine pre-treatment histopathology slide^32^. From cell-nucleus centroid point patterns extracted from H&E-stained slides, we jointly fit the 2PCF in Euclidean space and its corresponding power spectral density (PSD) in Fourier space to their analytically derived counterparts from the reaction-diffusion model. This joint calibration yields patient-specific effective net proliferation rate *γ* and diffusion coefficient D with high goodness-of-fit and practical identifiability across independent multicentric cohorts. From these primary parameters, we derive two pathology-motivated composites: the dispersion length, 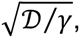 and the front velocity, 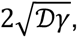 that link microscopic tumor tissue architecture to macroscopic tumor growth dynamics and carry prognostic information beyond standard staging.

Conceptually, HDTD is analogous to dynamical RNA-velocity methods such as scVelo, which infer gene-specific transcription, splicing, and degradation rates from static single-cell transcriptomes by fitting an explicit dynamical model^33^. Inferring proliferation and diffusion rates of solid tumors from histologic snapshots enables us to turn routine H&E-stained slides into a low-dimensional set of interpretable, mechanistic phenotypes. Since H&E imaging is universally and globally available across tumor types and health-care systems, it provides a pragmatic, pan-cancer, low-barrier approach to obtaining mechanistic biomarkers for forward simulations rooted directly in tissue architecture prior to treatment and immediately available at diagnosis without any change to the clinical workflow.

We applied HDTD to eleven independent cohorts that together comprise thousands of slides across a diverse group of solid human cancers. The calibrated parameters robustly reproduced spatial statistics of the observed patterns and were identifiable at the single-patient level. We employed these parameters to construct pan-cancer and intra-cohort maps of phenotypes, to stratify progression-free survival (PFS) and overall survival (OS), and to define a histology-calibrated mechanistic forward-simulation model. The utility of this model was demonstrated by rationalizing radiotherapy response from observed phenotypes in glioblastoma through an illustrative counterfactual simulation, which served as a full proof-of-concept pipeline. The HDTD framework may advance precision oncology by linking routine histopathology, mechanistic modeling, and model calibration to improve diagnosis and treatment planning.

## Results

### Reaction-Diffusion Parameters Can be Identified from Histopathology

For each patient, we extracted cell nuclei point patterns from H&E-stained slides, determined the corresponding 2PCF and PSD, and jointly fitted the kinetic parameters *γ* and D (see Methods section) for all regions of interest (ROIs) (Figure 1a). The cohort-level heterogeneities of identified 2PCF and PSD values in the head and neck squamous cell carcinoma (HNSCC) cohort are shown in Figure 1b as a representative example. The joint calibration achieved high fidelity across ROIs and datasets (2PCF: 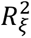 = 0.983 ± 0.085, PSD: 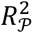 = 0.999 ± 0.005; mean ± standard deviation), resulting in an overall unweighted coefficient of determination *R*^2^ = 0.991 ± 0.045. The corresponding average normalized root mean squared errors (NRMSEs) for joint fitting on the 2PCF and the PSD were identified as *NRMSE*_£_ = 0.041 ± 0.028 and *NRMSE_P_* = 0.010 ± 0.009, respectively. We display two representative joint fits in Figure 1c. The joint fitting approach numerically stabilizes previous attempts to fit the PSD alone^34^ in terms of parameter identifiability and ROI edge correction. Profile likelihoods showed sharp minima and narrow confidence intervals for both parameters over all cohorts, and the 95% profile likelihood widths on average decreased from 0.39% ± 0.89% to 0.04 ± 0.39% (paired Wilcoxon signed-rank p<0.001) and from 4.73% ± 1.22% to 0.12% ± 1.28% (paired Wilcoxon signed-rank p<0.001) for the profiles of the fitted parameters *γ* and D, respectively (Supplementary Figure S1).

**Figure 1:**
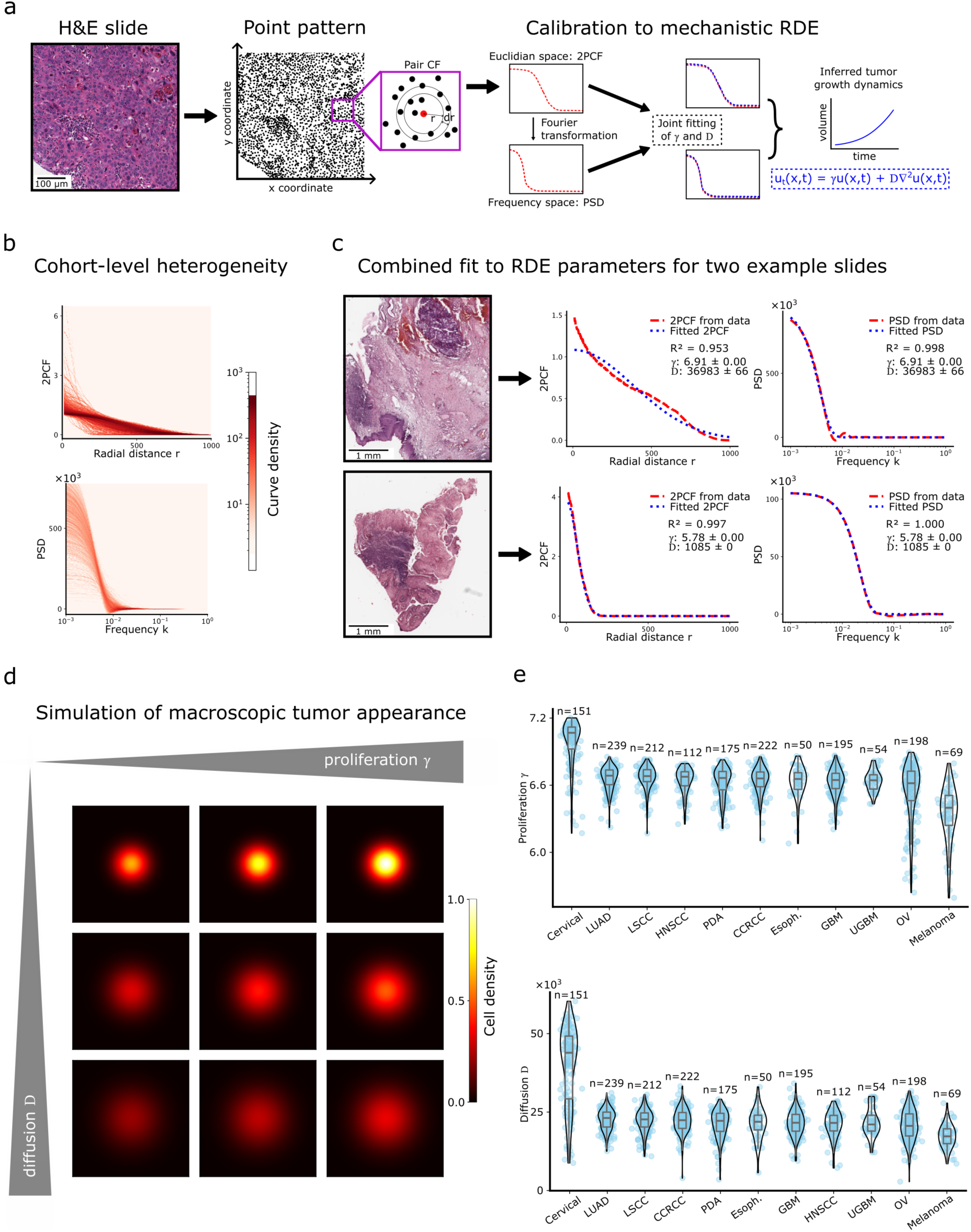
Calibration of reaction-diffusion dynamics from histopathology. Nucleus segmentation was performed on tumor tissue on routine H&E slides to derive centroid point patterns. The isotropic two-point correlation function (2PCF) in the Euclidean domain and the power spectral density (PSD) in the frequency domain, linked by Hankel transformation (Fourier transformation), are calculated from the pair correlation function (Pair CF). Joint fitting is performed on both functions to their analytically derived counterparts from the reaction-diffusion equation (RDE), which enables analytical formulation of tumor growth dynamics **(a)**. As a representative example, the cohort-level heterogeneities of identified 2PCFs and PSDs are illustrated for the HNSCC cohort **(b)**. Two representative joint fits of respective 2PCFs and PSDs to the RDE-derived analytical functions with reported identified parameter values and fit performances are illustrated in **(c)**. Macroscopic tumor appearance on forward simulations of the RDE are shown in **(d)**: Increasing the proliferation enhances overall amplification, whereas increasing diffusion smooths the spatial structure and yields coarser densities. The calibrated proliferation rate *γ* and diffusion coefficient D across the different cancers and cohorts are shown in **(e)** as box-violin plots to describe the respective cohort-level distributions. Comparisons of effective parameter distributions across cohorts are limited due to time normalization (see Methods).

### Parameters Define Mechanisms to Simulate Macroscopic *in silico* Tumors

The identified parameter space was utilized to forward simulate the mechanistic reaction-diffusion equation. Numerical simulations demonstrated that the macroscopic tumor phenotypes are defined by their respective parameter-dependent density fields (Figure 1d). This provides a biological explanation for the model parameters and their linked phenotypes and thus enables interpretation. Tumors with a highly proliferative (high *γ*) but low diffusive (low *D*) phenotype exhibit a dense macroscopic architecture. In contrast, tumors with a highly diffusive but less proliferative phenotype (low *γ*, high *D*) have more widely dispersed macroscopic structures. Pan-cancer analysis of the distribution of patient-specific (*γ*, *D*) parameter pairs demonstrates high variation within and across individual cancer types (Figure 1e).

### Point Process Simulations of Pan-Cancer Mechanistic Landscapes Recapitulate Diverse Established Morphological Architecture

Analogous to the continuous differential equation simulations, we evaluated discrete cell population morphologies from Thomas point process simulations with different (*γ*, D) parameter pairs. Higher diffusion parameters yielded a more dispersed architecture but with distinct differences in dense cluster formation within the established tumor and in the dispersive reach. Dispersion length, 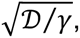 controls the spatial blur of tumor-cell clusters, whereas the front velocity, 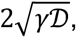 captures the growth-spread balance (Figure 2a). Low diffusion (D) coupled with high proliferation (*γ*) (low dispersion length) resulted in a more solid, high-grade microscopic appearance and heterogeneously distributed cell clusters. In contrast, high diffusion and low proliferation (low front velocity) yielded a more homogeneous, low-grade microscopic morphology where cell clusters are rare and more diffuse. The derived point-process morphologies are observed in pan-cancer patient tissue samples across different dispersion lengths (Figure 2b-e). Of interest, the H&E-stained slides from two patients with infiltrative pancreatic adenocarcinoma show different dispersion lengths (high-grade appearance with a derived low diffusion rate D and low-grade appearance with qualitative epithelial-mesenchymal transition-like structures and small cell groups with a derived high diffusion coefficient D) (Figure 2e). This suggests that the diffusion coefficient alone does not determine phenotypic invasiveness and that phenotypically comparable tissue slides may have different underlying mechanistic dynamics. Analyses of pan-cancer dispersion lengths and front velocities demonstrated high intra- and inter-cancer type heterogeneity (Figure 2f).

**Figure 2:**
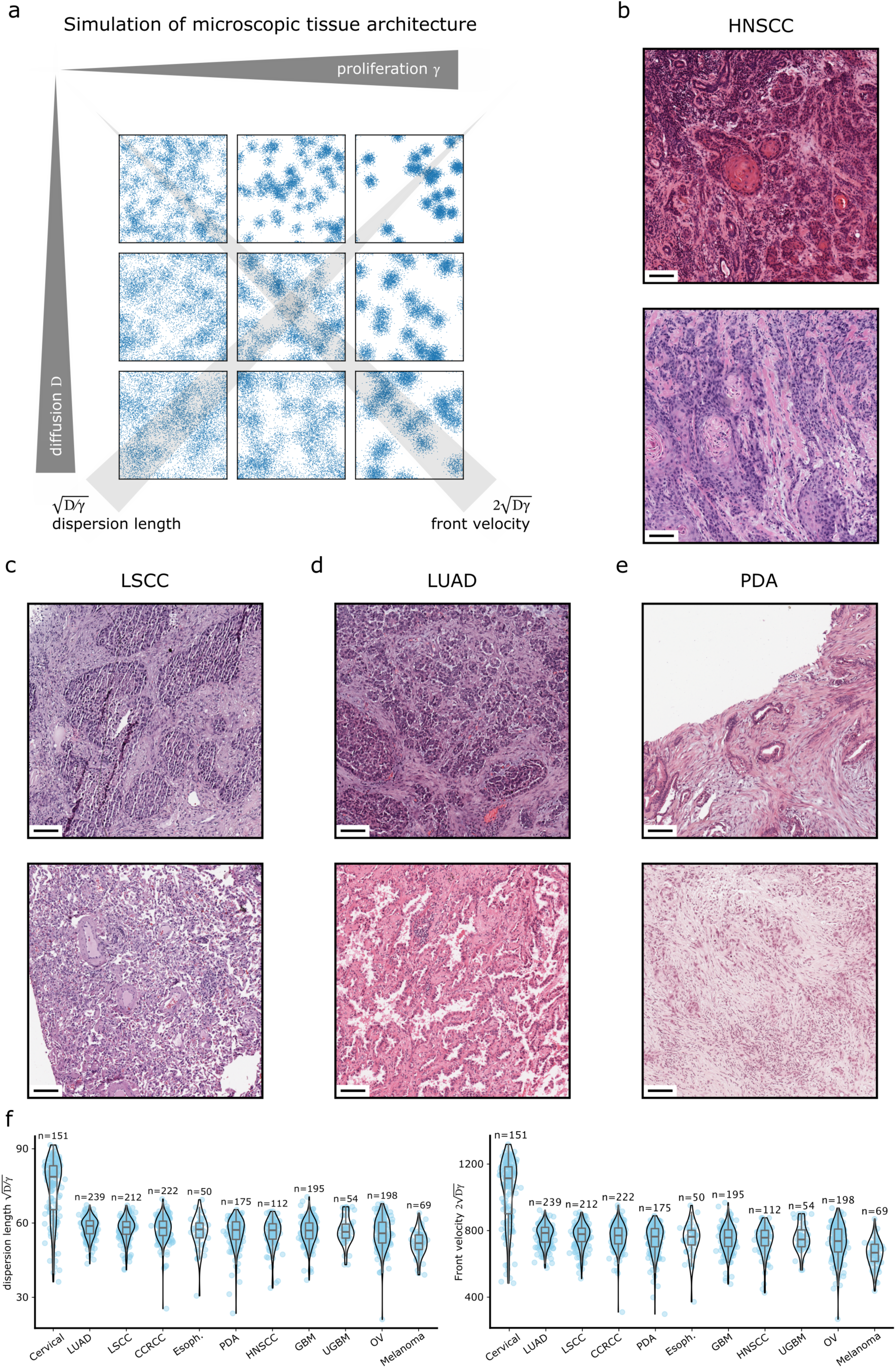
Diversity of mechanistic phenotypes and tissue architecture. Simulated microscopic tissue architectures for a grid of kinetic proliferation (*γ*) and diffusion (D) parameters illustrate how the reaction-diffusion parameters shape spatial clustering and dispersion of cells **(a)**. High proliferative and less diffusive phenotypes result in locally dense and heterogeneous microscopic architecture. Phenotypes characterized by low proliferation and high diffusion result in much more disperse and loosely connected microscopic tissue architectures. Panels **(b)**, **(c)**, **(d)**, and **(e)** show representative examples of H&E-stained tissue specimens from the head and neck squamous cell carcinoma (HNSCC), lung squamous cell carcinoma (LSCC), lung adenocarcinoma (LUAD), and pancreatic ductal adenocarcinoma (PDA) datasets to illustrate low diffusion/high proliferation regions (top row); these appear as dense, crowded cell clusters with relatively sharp boundaries in heterogeneous and high-grade tissues. The bottom rows of these panels show high diffusion/low proliferation phenotypes from the same dataset and exhibit more spatially dispersed nuclei, larger voids, and loosely connected structures in low-grade settings. H&E-stained tissues are shown at the same scale (bar indicates 100 µm). Panels were selected to illustrate morphological extremes of the joint parameter space. Of particular interest are the comparisons for pancreatic ductal adenocarcinoma in panel **(e)**: Both the low diffusion/ high proliferation (top row) and high diffusion/low proliferation (bottom row) samples show invasive traits. Thus, diffusion phenotypes do not necessarily translate into histologically apparent invasiveness at the sampled time point. The calibrated dispersion length 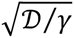 and front velocity 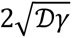 across different cancers and cohorts are shown as box-violin plots to describe the respective cohort-level distributions **(f)**.

### Parameters Reveal Pan-Cancer and Intra-Cohort Diversity of Mechanistic Phenotypes

The pair of calibrated parameters (*γ* and D) is referred to as a mechanistic phenotype that represents a quantitative trait of the tumor’s growth and spread behavior. The time normalization used to identify these kinetic parameters (see Methods) limits the direct comparison of identified parameters across cohorts. Thus, to investigate the differences on a common scale, we z-transformed the parameter values *γ* and D relative to the unweighted pan-cancer means and standard deviations (i.e., each parameter was standardized to a zero mean and unit variance across the entire cohort). The resulting pan-cancer parameter map on orthogonal trait axes *x* = *z_γ_* + *z_D_* and *y* = *z_D_* − *z_γ_* provides a compact, two-dimensional overview of the parameter landscape. The individual locations of parameter pairs within the parameter space differed from the overall unweighted averages of the datasets. Some tumor types occupied a prominent combined phenotype with some propensity for diffusion (cervical cancer), while others feature less pronounced combined phenotypes and had a propensity for proliferation (for example, head and neck or esophageal cancer), highlighting pan-cancer heterogeneity in proliferation and diffusion traits (Figure 3a). Additionally, we generated corresponding within-cohort parameter maps to visualize intra-cohort phenotype patterns that are not immediately apparent on the pan-cancer map. For this, we z-transformed the parameters analogously, but this time only for the parameter means and standard deviations for values within the same cohorts (Supplementary Figure S2). The resulting maps show within-cohort distributions of inferred growth-diffusion trade-offs and highlight the intra-cohort heterogeneities.

**Figure 3:**
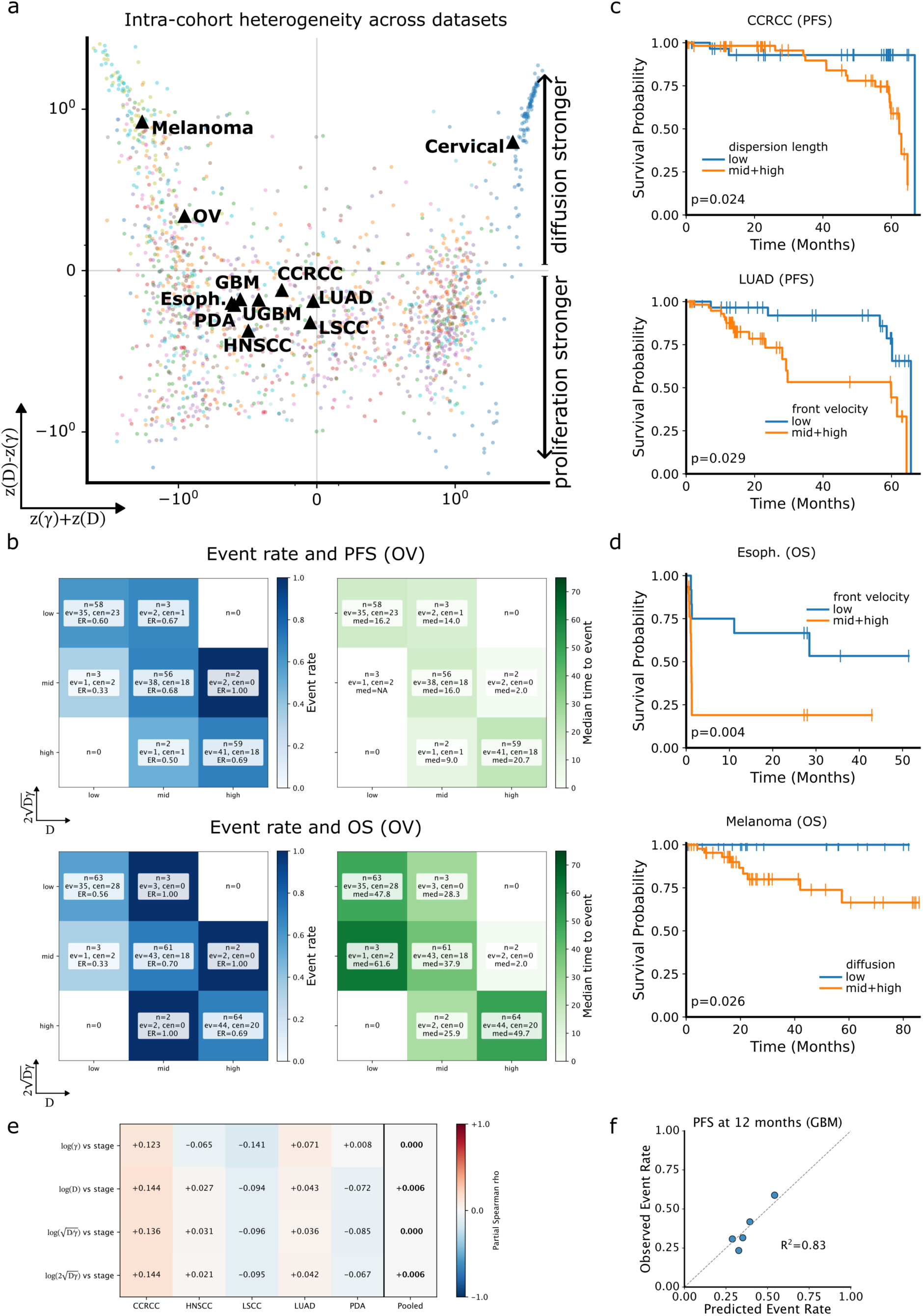
Pan-cancer reaction-diffusion phenotypes, intra-cohort risk, and histology-calibrated forward-simulations. Pan-cancer map using the z-transformed *γ*-*D* parameter space relative to global unweighted mean and standard deviations over all cohorts (**a**). Axes indicate directions of phenotype strengths: *x*-axis (defined by *x* = *z_γ_* + *z*_D_) reflects combined parameter strength; *y*-axis (defined by *y* = *z_D_* − *z_γ_*) separates diffusion- and proliferation-dominated phenotypes. Dots indicate patient-specific parameters, color-coded by dataset. Black triangles indicate cohort centroids and demonstrate inter-cohort heterogeneity. Event rates (left) and Kaplan-Meier-estimated median time to event (right) are shown for each phenotype for PFS (top) and OS (bottom) in the ovarian cancer cohort **(b)**. Nine phenotypes are defined by tertiles of the parameters D and 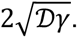 Tiles describe the number of patients (n), events (ev), censored patients (cen), event rate (ER), and median time to event (med). For OS and PFS Kaplan-Meier tile estimations showed a global log-rank *p* < 0.0001. Analogous plots for all other cohorts are shown in Supplementary Figures S3-S6. Stratification for intra-cohort phenotypes as defined by parameter distribution tertiles shows Kaplan-Meier survival curves for PFS in the clear cell renal cell carcinoma (CCRCC) and the lung adenocarcinoma (LUAD) cohorts **(c)**, and for OS in the esophageal (Esoph.) and melanoma cohorts **(d)**. Statistical test: log rank test. Orthogonality of HDTD parameters to anatomical stage across five CPTAC cohorts with standardized TNM staging **(e)**. The heatmap shows partial Spearman rank correlations of log-transformed HDTD pairs (log *γ*, log D) and 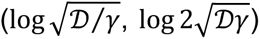 with ordinal stage, residualized on age and sex. The rightmost column displays Fisher-z pooled estimates. All pooled absolute correlations are below 0.01, and between-cohort heterogeneity is not detectable (Cochran *Q p* ≥ 0.30, *I*² between 0.04 and 0.17). sBM lacks standardized TNM staging and is excluded. Calibration of the CPH model (*γ*, D) at 12 months for the CPTAC glioblastoma cohort **(f)**. Patients are binned into five equal-sized groups by predicted event probability. Mean predicted event rates per bin are plotted against observed event rates for PFS, dashed lines indicate perfect calibration.

### Progression-Free and Overall Survival for Intra-Cohort Mechanistic Phenotypes

Patients were stratified by mechanistic phenotypes, defined as tiles in the two-dimensional parameter combination spaces. Within each cohort, the tiles corresponded to joint quantiles of the two parameter dimensions, and survival differences between tiles were assessed using Kaplan-Meier curves and log-rank tests.

The 3×3 tiling in the 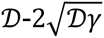 plane identified phenotypic differences in diffusion and front velocity to be associated with ovarian cancer PFS (global log rank *p* < 0.0001) and OS (global log rank *p* < 0.0001) (Figure 3b). Pan-cancer tiling and phenotypic variations, where survival data were available, are shown in Supplementary Figures S3 and S4 (PFS) and S5 and S6 (OS). Kaplan-Meier curves showed a difference in PFS for low versus intermediate/high front velocity in the clear cell renal cell and lung adenocarcinoma cohort (log rank *p* = 0.024 and *p* = 0.029, Figure 3c). Furthermore, OS differed significantly between low and intermediate/high front velocity in the esophageal cohort and between low and intermediate/high diffusion in the melanoma cohort (log rank *p* = 0.004 and *p* = 0.026, respectively; Figure 3d). Pan-cancer trends in PFS curves are shown in Supplementary Figure S7.

### HDTD Parameters are Orthogonal to Anatomical Stage across CPTAC Cohorts and add Prognostic Information in Glioblastoma

Partial Spearman rank correlations between each HDTD parameter and ordinal stage, residualized on age and sex, were calculated across the five CPTAC cohorts with standardized TNM staging (CCRCC, HNSCC, LSCC, LUAD, PDA) and were small per cohort (maximum absolute value 0.124). Fisher-z pooling with inverse-variance weighting yielded a pooled absolute correlation below 0.01 for every HDTD parameter (Figure 3e), and between-cohort heterogeneity was not detectable, with Cochran’s *Q p* ≥ 0.30 for every parameter and *I*^2^ between 4% and 17%. HDTD parameters capture a mechanistic axis that is statistically independent of anatomical stage. GBM was excluded in this sub-analysis due to unavailable TNM stage. A Cox proportional-hazards (CPH) baseline model (Age, Sex) was fitted in the glioblastoma cohort, and compared against an augmented CPH model (Age, Sex, log-transformed primary HDTD pair (log *γ*, log D)). For PFS (n=87, 52 events), the augmented model increased Harrell’s c-index from 0.57 to 0.62 (absolute increase 0.05, relative increase from chance level 69.6%; likelihood-ratio *p* = 0.036). Continuous reclassification at 12 months yielded an IDI of 0.07 (95% CI [0.00, 0.20]) and NRI of 0.33 (95% CI [−0.11, +0.93]). Calibration for PFS at 12 months by five equal-sized predicted-risk bins followed the identity line (*R*^2^ = 0.83, Figure 3f), and Schoenfeld residual tests showed no proportional-hazards violation (minimum covariate-level *p* = 0.720). A higher effective kinetic proliferation parameter *γ* was associated with longer PFS (HR 0.73, 95% CI [0.56, 0.96], *p* = 0.024), cross-validated (fivefold, out-of-fold c-index gain 0.04, 95% CI [0.01, 0.07]) and attenuated but directionally unchanged after adjustment for age and sex (HR 0.69, 95% CI [0.52, 0.91]), IDH mutation status (HR 0.76, 95% CI [0.58, 1.01]) and gene-level MGMT methylation (HR 0.77, 95% CI [0.59, 1.01]). Along molecular signatures measured from the same tumors in the GBM cohort with mass-spectrometry proteomics, a six-protein proliferation signature (MKI67, TOP2A, CCNB1, RRM2, PCNA and CDK1 with Cronbach’s alpha 0.955) correlates with the effective proliferation parameter at only 0.14 (95% CI [−0.06, +0.33]). None of the six proteomic axes spanning mesenchymal transition, hypoxia, angiogenesis, myeloid infiltration, stemness and proliferation reaches nominal significance against either parameter. The proliferation signature alone was not found to carry prognostic information in this cohort (HR 1.06, 95% CI [0.84, 1.34], *p* = 0.603), whereas in a cross-validation the single histology-derived parameter *γ* alone carries more prognostic information than the full 41-protein panel (out-of-fold c-index 0.61 ± 0.01 versus 0.58 ± 0.03) and it preserves its magnitude after adjustment for the proliferation signature and the six proteomic axes measured on the same tumors (HR 0.74, 95% CI [0.55, 1.00], *p* = 0.052), against an unadjusted HR 0.73 (95% CI [0.56, 0.96], *p* = 0.024) on the same patients.

### Histology-Calibrated Forward Simulations Provide an Explainability Layer for Radiotherapy Response and Identify Favorable Proliferative Phenotypes

The potential applications of histology-calibrated forward simulations and treatment responses inferred from growth dynamics were illustrated as proof of concept with alternative radiotherapy schemes in glioblastoma. Kaplan-Meier curves for proliferation *γ* split at the median level showed a significant difference in PFS between the lower and higher proliferation groups (log-rank *p* = 0.023, Figure 4a). Using four representative parameter combinations from the glioblastoma cohort (high proliferation, high diffusion, etc.; cf. Figure 1d), we simulated radially symmetrical tumor growth to a fixed macroscopic diameter to mimic comparable clinical presentations for the different phenotypes, shown as cell density fields and radial density profiles (Figure 4b).

**Figure 4:**
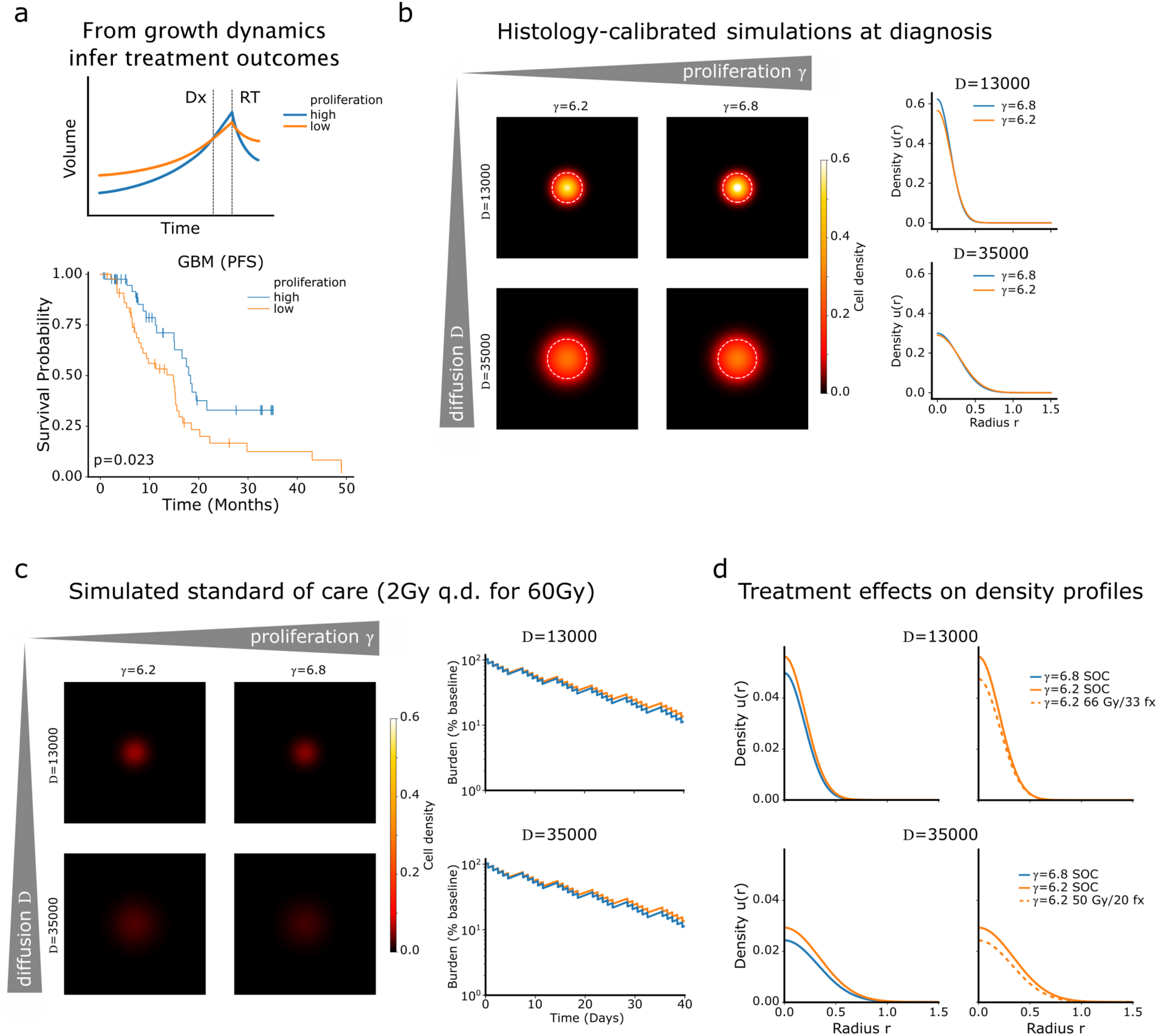
Histology-calibrated mechanisms provide explainability layer for radiotherapy response of inferred phenotypes in GBM, and forward simulations enable patient-specific digital twins and counterfactuals. Two tumors of the same size at diagnosis differ in their pre-diagnostic growth and treatment response. Growth dynamics inferred from histopathology explain the underlying biology: for the glioblastoma (GBM) cohort split at the median inferred proliferation rate, the high proliferation group shows longer progression-free survival (*p* = 0.023, log-rank test). This is consistent with linear-quadratic radiobiology (cf. Methods) **(a)**. Histology-calibrated simulations of four phenotypes grown to the same macroscopic size at diagnosis (dashed circle) shown as cell density fields in cartesian and density profiles in radial dimensions **(b)**. After simulated standard of care (60 Gy in 30 fractions, 2 Gy daily) every phenotype falls below detection limit, but the residual burden is different with 11.0% of the baseline for high proliferation and 13.3% of the baseline for the low proliferation phenotypes. The forward simulations allow to investigate the total burden over the treatment window **(c)**. Under identical standard course, the low proliferation branch (orange) maintains a higher residual burden than the high proliferation branch (blue). Dose-escalating schedules can be characterized with the model to map the former onto the latter (cf. Methods). For low diffusion 66 Gy in 33 fractions (2 Gy per fraction) and for high diffusion 50 Gy in 20 fractions (2.5 Gy per fraction) were identified **(d)**.

Under simulated standard of care radiotherapy (2 Gy once daily to 60 Gy in 30 fractions, i.e., six weeks of treatment) all simulations post-treatment showed tumor densities below the assumed clinically detectable threshold. The total tumor burden was reduced to 11.0% of its baseline value for high proliferation phenotypes and to 13.3% for the low-proliferation phenotypes (Figure 4c). The phenotype with higher proliferation ends treatment with the lower residual burden, since the linear-quadratic treatment effect links radiosensitivity to the proliferation rate. This is consistent with the longer PFS for the high proliferation group. In addition, the forward simulations allow to ask which schedule applied to the low proliferation phenotype reproduces the post-treatment density profile of the high proliferation group after standard of care. We found this to be dependent on the diffusion coefficient: For low proliferation and low diffusion, the schedule reproducing the mentioned radial density profile was found to be a moderate dose intensification to 66 Gy in 33 fractions (2 Gy per fraction), whereas for low proliferation and high diffusion, the schedule was 50 Gy in 20 fractions (2.5 Gy per fraction) (Figure 4d).

### Parameter-Stratified Virtual Trial Enables Counterfactual Testing of Treatment Design

The comparison above fixes the time points but not the outcome. To address what the inferred kinetics can report at a trial level, we converted each patient’s simulated schedule contrast into a shift of their observed event time and re-censored such that the observed survival curve under standard of care serves as the reference arm and every alternative schedule is a counterfactual simulation on the identical patients^35^. Six candidate schedules were considered: 75 Gy in 30, 70 Gy in 35, 60 Gy in 30, 50 Gy in 20, 40 Gy in 15 and 30 Gy in 5 fractions. All 87 patients with a published progression-free survival (52 recorded events) were enrolled in the virtual trials. Counterfactual simulations that give a deescalation compared to the standard of care had to be restricted to 78 patients (44 events) to ensure that the counterfactual event time is kept as positive and thus logically feasible. Unlike the phenotype simulations above, this comparison evaluates the schedule contrast in closed form and enters only the calibrated proliferation rate (see Methods). The virtual cohort as parameter distribution on the absolute proliferation *γ*^∗^ was calibrated on the standard of care, and the observed PFS curve from Figure 4a was exactly recovered with a median PFS 18.3 versus 14.8 months (log-rank *p* = 0.023, Figure 5a). As a first counterfactual, we simulated the dose-intensified arm of the NRG-BN001 trial^36^, 75 Gy in 30 fractions (2.5 Gy per fraction) delivered within the same six weeks as standard of care, assigned to each patient. Since the number of treatment days did not change, the protraction term canceled out, and all counterfactual times stayed positive. Thus, all 87 patients are eligible for forward simulations for this virtual trial. The restricted mean PFS rose by 3.5 months (95% CI [2.5, 4.6]) and the median PFS by 5.0 months, and the gain was found to be present mostly in the low proliferation stratum (5.4 versus 1.1 months RMST at 24 months for low and high proliferation, Figure 5b). The treatment benefit was investigated depending on the value of the absolute proliferation *γ*^∗^ through forward simulations of the different available schedules. Some heterogeneity was found for the days gained depending on schedule and parameter (Figure 5c) and we constructed a virtual personalized clinical trial by splitting the cohort at the median level of the effective proliferation parameter *γ*, and adjusting each branch to opposite directions: the slower proliferating half received the escalating course while the faster proliferating half received the deescalated course. The two branches were then pooled back into a single population curve. 43 patients above the median were allocated to 30 Gy in 5 fractions and 44 below it to 75 Gy in 30 fractions, i.e., faster growing tumors to the shorter course with the higher dose per fraction and slower tumors to the dose-escalation schedule (Figure 5d). Within the low-proliferation group, every deescalated schedule performed worse than the standard of care, by 18.7, 48.7 and 66.9 days of restricted mean survival for 50 Gy in 20, 30 Gy in 5 and 40 Gy in 15 fractions respectively, in contrast to the high-proliferation group where 30 Gy in 5 fractions was better by 17.8 days and 50 Gy in 20 fractions by 6.8 days, while 40 Gy in 15 fractions was 1.8 days worse. None of the deescalated uniform schedules improved over the standard of care: given to everyone, 50 Gy in 20, 30 Gy in 5 and 40 Gy in 15 fractions lost 5.0, 11.8 and 30.7 days of restricted mean survival at 24 months, and 4.0, 10.1 and 18.4 days at 12 months. The dose-intensified 70 Gy in 35 and 75 Gy in 30 fractions gained 25.4 and 69.6 days at 24 months (12.1 and 27.0 days at 12 months). Against standard of care (60 Gy in 30 fractions) the pooled population curve for the stratified treatment design gained 3.2 months (95% CI [2.3, 4.2]) of RMST at 24 months. The median PFS rose from 15.2 to 19.0 months (Figure 5d). No log-rank test is reported since the compared arms are not independent. Instead, the schedule mix was kept fixed, and the patient assignment was randomly assigned to test whether *γ* carries no allocation information. None of the 2,000 random assignments reached the observed difference, and a random assignment to these two schedules gained 26.4 ± 7.5 days against the standard of care (permutation *p* < 0.001). For the deescalation-only design the corresponding control lost 6.6 ± 2.7 days against the standard of care (permutation *p* < 0.001). We conclude that the difference lies in which patient receives which course in the stratified manner rather than in a random assignment of schedules.

**Figure 5:**
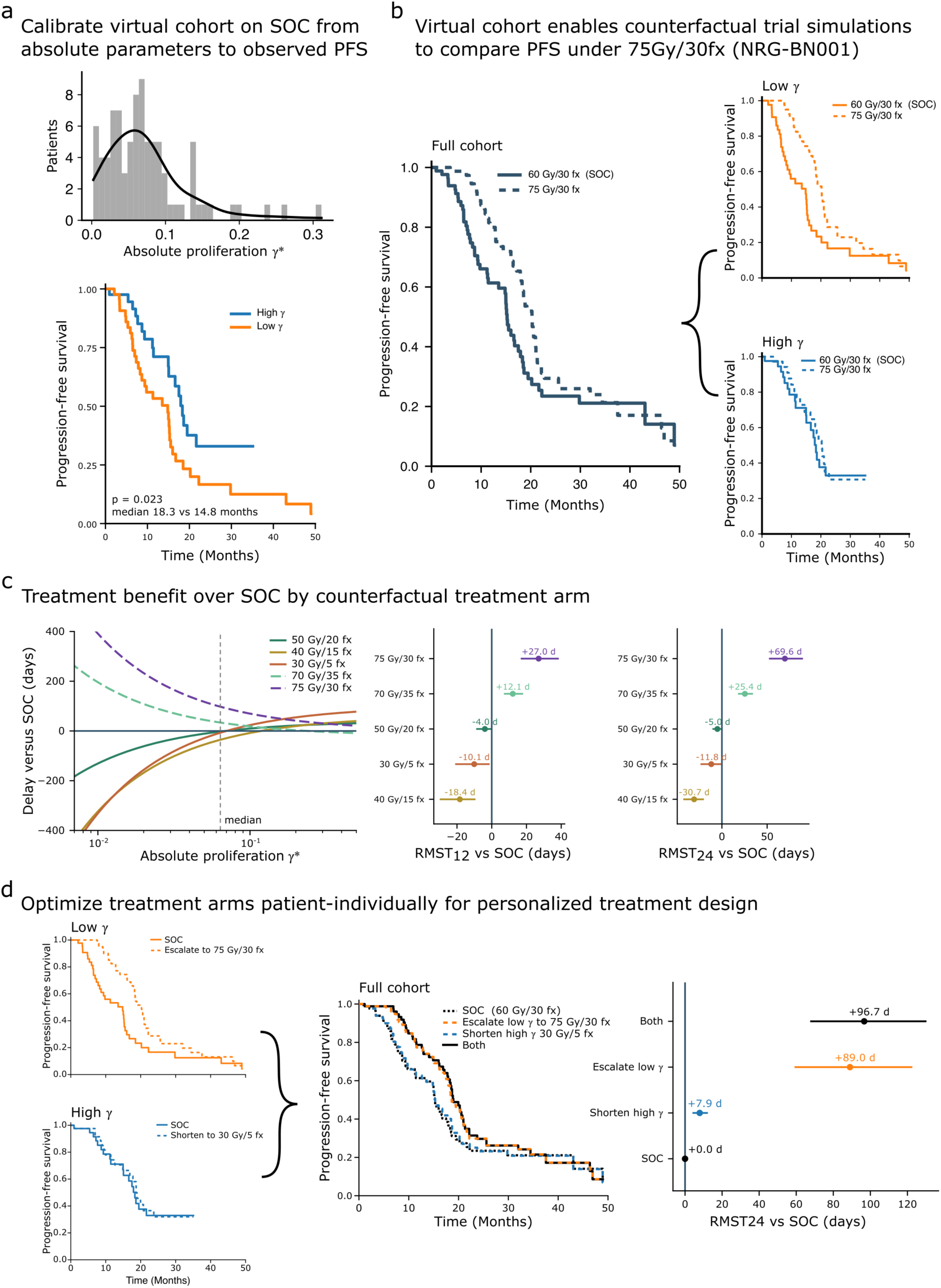
Virtual trials of histology-calibrated digital twins identify why trials fail and enable counterfactual trial simulation and patient-individual treatment design. The virtual cohort is defined by the decompressed patient-specific absolute proliferation rates of all 87 patients with GBM with published progression-free survival (PFS; cf. Methods). The calibration on standard of care (SOC) reproduces the observed PFS including the median split **(a)**. The dose-escalating arm of the NRG-BN001 trial with 75 Gy in 30 fractions (2.5 Gy per fraction) is assigned to each virtual patient, increasing median PFS over SOC by 5.0 months on the population level. Stratifying for the identified proliferation parameters at the median, the comparison of survival curves uncovers that the benefit sits in the slow proliferation branch at a median PFS increase of 5.4 months versus 1.9 months for the fast proliferation branch **(b)**. No log-rank test can be performed for these comparisons since the two arms are not independent. The treatment benefit over SOC is shown as a function of the absolute proliferation rate for each uniform schedule, and as the restricted mean survival times (RMST) at 12 and 24 months with 95% bootstrap intervals (cf. Methods). This shows a heterogeneity in benefits based on the identified parameter and chosen schedule **(c)**. Stratum-specific reassignment against SOC escalating the slow proliferation branch to 75 Gy in 30 fractions (+3.0 months, [2.0, 4.1]), shortening the fast proliferation branch to 30 Gy in 5 fractions (+0.3 months, [0.1, 0.4]) and the stratified design combining the two (+3.2 months, [2.3, 4.3]). The plots show the benefits in PFS for the two branches and the full population, as well as the added RMST at 24 months compared to SOC with 95% bootstrap intervals **(d)**.

## Discussion

We show that patient-specific kinetic parameters of a mechanistic reaction-diffusion equation (RDE) can be inferred directly from a single routine H&E-stained histopathologic slide, and that these parameters carry prognostic information beyond standard staging in glioblastoma. We combined spatial point pattern analysis with analytical solutions derived from the RDE by jointly fitting the 2PCF and the PSD to obtain an effective net proliferation rate and diffusion coefficient for each tumor. From these, we were able to determine the mathematically motivated dispersion length and tumor front velocity to further characterize each tumor. These four metrics define a low-dimensional space and a mechanistically interpretable perspective of tumor phenotypes that can be estimated from a single slide.

In diffuse glioma, MRI-calibrated RDEs have been used to estimate patient-specific proliferation and diffusion rates to predict growth and recurrence and to inform radiotherapy design or other treatment strategies^15,18,37,38^. Our work expands mechanistic reaction-diffusion modeling in oncology from radiology image-based calibration in a subset of cancers to a much broader histology-based setting. We have demonstrated that analogous kinetic parameters can be inferred from the microarchitecture of routinely collected tissue samples from multiple types of solid tumors. This complements current efforts in computational and digital pathology to predict molecular alterations, immune phenotypes, and outcomes based on histopathology using deep neural networks^1–4^. Unlike purely data-driven models, our framework yields kinetic parameters that are directly embedded in an explicitly formulated dynamical system with a clear biological and mechanistic interpretation. This type of model allows for the generation of patient-specific forward-simulation models, where patient-specific counterfactual treatments can be simulated in real time to guide individual clinical decision-making^39–41^.

Our results demonstrate that data analyses from the 2PCF in Euclidean space and the PSD in Fourier space combined characterize an empirical correlation signal represented in two linked domains to derive RDE dynamics from a static histopathology slide. This complements other recent attempts to derive cell-population dynamics from a single spatial proteomics snapshot^31^ with relevant clinical actionability. HDTD differs in two aspects relevant for clinical deployment: it operates on H&E histopathology, which is performed for nearly every solid-tumor patient worldwide and does not require specialized molecular assays, and it calibrates an explicit reaction-diffusion partial differential equation with closed-form analytical solutions that can be directly embedded in forward simulations of the calibrated model. Previous work has shown that spectral analysis of biopsy tissues can be used to calibrate simplified tumor growth and invasion models from histology^34^, but these studies did not use joint fitting, which we have shown to be superior in fit performance and stable in parameter identifiability. Joint fitting substantially narrows the profile likelihood confidence intervals compared to PSD-only calibration. Together with the high goodness-of-fit across the heterogeneous cohorts, this suggests that the RDE model captures the dominant length scales and correlation structures of the tumor cell organization.

We used microscopic and macroscopic simulations to explore the derived phenotypes. At the microscopic scale, different combinations of *γ* and D manifest as dense, high-grade versus dense, low-grade architectures. These regimens were illustrated with representative histological examples across cancer types. At the macroscopic level, we used the calibrated parameters in forward simulations of the mechanistic RDE to visualize their effects on tumor compactness, dispersion, and front propagation. These results connect cell-scale architecture to imaging-scale growth behavior and align with previous work linking spatial organization in histopathology and multiplex imaging to functional states of the tumor microenvironment^28^.

We found that the calibrated parameters were not only interpretable but also clinically applicable. Across several cohorts, specific regions on the parameter map were stratified by PFS and OS. Stratification of diffusion and front velocity parameter planes and of single parameters yielded consistent separation of Kaplan-Meier survival curves in glioblastoma, pancreatic cancer, ovarian cancer, lung adenocarcinoma, clear cell renal cell carcinoma, esophageal cancer, and melanoma. These findings complement established deep learning-based survival models operating in high-dimensional feature spaces^4,7^ and suggest that a small number of mechanistic parameters can capture prognostically relevant tumor dynamics. Associations varied across cohorts, reflecting differences in patient, tumor, and treatment characteristics. The identified parameters should be viewed as complementary mechanistic descriptions rather than replace established molecular and histological markers.

The identified parameters are calibrated against a mechanistic dynamical model and can therefore serve as the mechanistic core component of mechanistic forward simulations. In a counterfactual comparison the cohort was stratified by the inferred proliferation parameter, and each arm received a different established radiation schedule. The faster growing branch was simulated to receive a one-week hypofractionated course, and the slower growing branch to receive a dose-escalating schedule on the same treatment duration. The resulting population curve exceeded standard of care and every single-schedule alternative *in silico*. It preserved most of the gains of uniform dose intensification while sparing the faster proliferating branch most of the dose and time on treatment. The reason for this is that the two stratified groups respond in opposite directions. The faster growing branch is relatively insensitive to dose deescalation, while the slower proliferating branch benefits from additional biologically effective dose. We demonstrated that histology-calibrated proliferation rates can yield distinct response dynamics, and that after identical standard treatment courses, the identified phenotypes show different residual burdens. In principle, mapping the post treatment density of the slower phenotypes onto the one of the faster phenotypes is possible, but requires treatment adjustments.

Recent conceptual and technical work on oncology digital twins emphasizes the need for parsimonious, clinically accessible model inputs^19,40,41^. HDTD directly addresses this requirement: it is designed to enable the conversion of raw routine H&E slides into mechanistic features for research and development pipelines. Image-derived kinetic parameters can serve as biomarkers for patient stratification in clinical trials by, for example, discriminating between rapidly growing, less diffusive tumors and more dispersive phenotypes that might require different treatment modalities, intensities, or combinations. The parameters are introduced as a continuous metric rather than a mere classification and can further be used in forward simulations to explore “what-if” scenarios for potential treatment avenues. They could also complement AI-based histology signatures and multi-omics or spatial-omics predictors, particularly when mechanistic interpretability is desired. Since the presented method only requires digitized slides and standard computing resources, it is compatible with existing digital pathology workflows and with retrospective pan-cancer databases. The presented counterfactual trial is a minimal instance of such a framework used for clinical decision-making. Its input are a digitized diagnostic slide and a published radiobiological constant, and its output is an allocation rule with a stated decision to a specific treatment design. This level of parsimony allows the mechanistic framework to easily enter clinical workflows and potentially serve as prior to define a digital twin, separating decision support tools from pure simulations that only reproduce observed behavior. In turn, our framework can put into perspective and be cross-checked with experience from real clinical trials. Radiotherapy is prescribed as a one-size-fits-all rule, and in a pooled pan-cancer analysis of 1,615 patients the derived biological effect of a delivered dose predicted the benefit of radiotherapy, while the physical dose did not^42^. This framework of a per-patient radiosensitivity index was independently prognostic in a glioblastoma cohort^43^. Glioblastoma further features an example of a patient-dependent treatment optimum: in the Nordic trial 34 Gy in 10 fractions outperformed 60 Gy in 30 fractions above the age of 70 (HR 0.59, *p* = 0.02), while the standard of care was appropriate for younger patients^44^. For patients aged 65 and above, the schedule for 40 Gy in 15 fractions became an accepted treatment schedule shortly afterwards^45^. The stratifiers we propose are of a mechanistic nature rather than demographic one, and whether an inferred proliferation rate stratifies better than age is a question only a prospective trial can answer. The dose-intensification has been addressed prospectively in the photon cohort of the NRG-BN001 trial^36^, though on a different endpoint. Our simulations suggest a progression-free benefit that is concordant in direction. We also want to underline that our results do not support that a 75 Gy in 30 fractions is superior to the standard of care 60 Gy in 30 fractions schedule as a single-arm treatment schedule, but that our results should rather be interpreted in the form that cohorts could be sub-stratified with mechanistic interpretable kinetic signatures to enhance patient-individual outcome rather than optimizing cohort-median outcomes.

The presented approach has several limitations. First, whole-tissue calibration could dilute or confound the results due to inclusion of non-relevant areas. Single-slide parameter estimates featured limited reliability across multiple slides of the same tumor (intraclass correlation 0.15), so patient-level values aggregate results from multiple regions. The method is, however, also directly applicable to selected sub-regions. Second, all nuclei within a region are treated as a single cell type, with shared kinetic parameters. Currently, tumor-immune-stroma interactions and cell-state heterogeneity, which are known to influence progression and treatment response^28,46^, are not differentiated. Extensions using multiplex immunofluorescence, immunohistochemistry, or AI-based cell-type classification to construct multiple point patterns and calibrate coupled RDE systems for interacting tumor, immune, and stromal populations could identify biological correlates of the kinetic parameters. Third, ROIs were determined using size constraints and downsampling enabling an automatic, scalable pipeline directly from H&E samples, which may bias the spatial statistics. Pre-defined ROIs within the slides to identify microscopic phenotypic differences (e.g., invasive fronts or low cellularity regions) with specific cell types should be tested. Fourth, the analyses were limited to two-dimensional tissues and their corresponding two-dimensional correlation functions. Extending HDTD to three-dimensional histology and time-resolved imaging could further capture complex tumor geometries and dynamics, particularly for anisotropic structures. Evidently, the recovered kinetic parameters are effective rates for an idealized isotropic reaction-diffusion mechanistic model rather than direct measures of cellular proliferation or motility. Consistent with this, the effective proliferation parameter correlates with the Ki-67 index (an absolute proliferation measure) only weakly in the glioblastoma cohort. Finally, the study is a retrospective and explorative evaluation of multicenter pan-cancer cohorts. Prospective validation studies will be required to establish robust generalizability and clinical utility.

The retrospective evidence presented in this work shows prognostic associations of the HDTD-derived parameters in several of the cohorts with available survival data. We deliberately do not over-claim the predictive validity in the sense of induced parameter-dependent treatment changes that would require formal interaction analyses against systemic treatment variations and that the retrospective data do not support. A directly testable use case is the fractionation choice in glioblastoma, for which the presented analysis specifies the design as a two-arm allocation of 30 Gy in 5 fractions for the faster proliferating and 75 Gy in 30 fractions for the slower proliferating half, split at the median of the effective proliferation parameter *γ*^∗^ showing improvements in PFS over the established standard of care in a virtual trial. What would need to be calibrated and validated before such a design could be run as an actual clinical trial is the link between the effective proliferation parameter and radiosensitivity, which cannot be estimated on a cohort in which every patient received the same schedule, particularly since the mechanistic agreement with a proteomic proliferation signature measured on the same tumors was found to only be 0.14. The parameter *γ* is coupled to the established linear-quadratic radiobiology model and could be evaluated against natural treatment variations that are contained in completed prospective trials. Further, prospective single-arm validation cohorts with a parallel mechanistic validation of *γ* against Ki-67 or mitotic-count data on paired H&E-stained biopsies would be the most informative subsequent experiments. Both are feasible without requiring changes to the clinical workflow.

Despite these limitations, our study shows promise for robustly inferring mechanistic RDE parameters from routine single-timepoint histopathology. The resulting phenotypes are interpretable, clinically informative in several settings, and directly utilizable for forward simulations. Thus, the presented method has the potential to shift the paradigm from descriptive pathology to mechanistically interpretable pathology. The derived parameters are effective net rates in the context of the chosen mathematical modeling approach and not translatable to other mathematical models with different functional forms and biological complexities. It is conceivable, however, that different mathematical approaches may be linked with analogous procedures for describing the metastatic process^47–49^ or treatment effects^50,51^ into multiscale patient-specific forward-simulation models^39^.

## Methods

### Calibration of Dynamics from Single-Timepoint Histopathology

Regions of interest (ROIs) were extracted from routine H&E-stained slides for each patient, the individual cell nuclei were segmented, and their coordinates were represented as two-dimensional point patterns. The spatial organization is characterized by the isotropic two-point correlation function (2PCF) in Euclidean space and the corresponding power spectral density (PSD) in Fourier space. The proliferation rate *γ* and the diffusion coefficient D of the mechanistic dynamical reaction-diffusion model are then calibrated by fitting the tissue 2PCF and PSD to the analytically derived reaction-diffusion equation (RDE). For each patient, maximum-likelihood estimates and profile likelihoods of *γ* and D are calculated to assess identifiability and uncertainty. If patients had multiple ROIs, the respective patient-level median of each value was used as the representative patient-specific metric for further analysis.

### Mechanistic Dynamical Reaction-Diffusion Model

The local density of cells *u*(*x*, *t*) at a given two-dimensional location *x* ∈ ℝ^2^ and at time *t* ∈ [0, ∞) was modeled using the reaction-diffusion partial differential equation

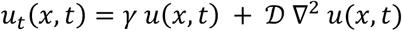

with initial condition *u*(*x*, 0) = *δ*(*x*). Here, the parameters *γ* > 0 and D > 0 denote the net proliferation [1/time] and the net dispersal rate [area/time] of cells, respectively. Two further major metrics for characterizing tumor densities simulated with this modeling approach are based on reaction-diffusion equations and borrowed from the field of mathematical biology^52^: the characteristic dispersion length 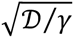 and the front velocity 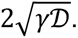 These biologically interpretable dynamics explain mechanistically how far the density moves relative to a proliferation turnover and how fast the propagation speed of the density’s outer rim is. The front velocity is the asymptotic travelling-wave velocity of the nonlinear Fisher-KPP equation. The linear equation above governs the leading edge of such fronts where density tends to zero, which justifies the use of the parameter pair calibrated from the linear regime to parameterize 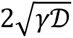 as a proxy for macroscopic propagation speed under density-limited growth. The same convention has been used previously in histology- and imaging-calibrated glioma modelling and allows for direct comparison^13,14,18,37,38^.

### Analytical Power Spectral Density and Two-Point Correlation Function

The analytical closed form for the PSD *P*(*k*) for a frequency *k* of the reaction-diffusion partial differential equation can be determined through Fourier transformation *u*U(*k*, *t*) of the equation’s solution *u*(*x*, *t*) with

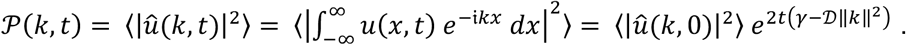

From the above-mentioned initial condition, we derive

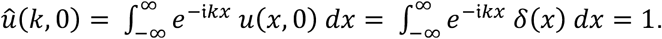

Therefore, the final analytical form for the PSD is

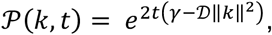

and following the Wiener-Khinchine theorem^53–56^, 2PCF is calculated as the inverse Fourier transformation of the PSD:

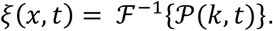

Assuming perfect isotropy, the 2PCF is reformulated in polar coordinates (here in two dimensions) as

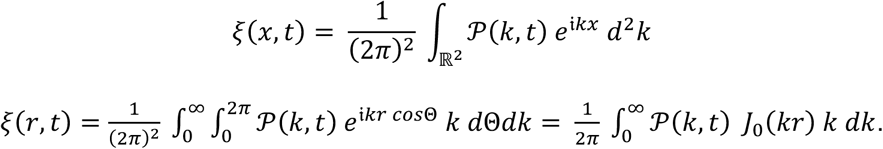

With the above PSD in two dimensions, we directly receive the analytical form

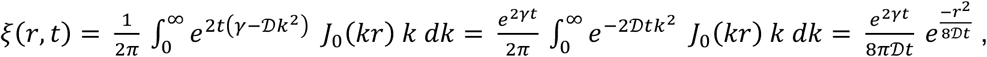

using Gradshteyn and Ryzhik’s Bessel identity^57^. For all analytical expressions of 2PCF and PSD, time was set to *t* = 1 to estimate *γ* and D. This normalized time scale allows for the interpretation of the parameters as effective net proliferation rate or diffusion coefficient in pseudo-time relative to the age of the tumor. The absolute units therefore depend on the choice of spatial and temporal scaling. Identification of time on an absolute time scale is, in principle, possible, but would require at least two observations of the same tissue to enable inference. We focused on a method that uses only one pre-treatment tissue sample and therefore followed the approach for relative time inference when estimating the parameters of the dynamical model.

### Histopathology Data

We retrospectively analyzed digitized pre-treatment whole-slide images and core needle biopsies from a diverse set of institutional and publicly available datasets (Table 1). The use of institutional datasets (melanoma, cervical cancer, esophageal cancer) was approved by The University of Texas MD Anderson Cancer Center Institutional Review Board (IRB 2023-0971, IRB 2025-0243, and IRB PA17-0191) under waived informed consent. Every step in the analysis (nucleus segmentation, clustering, spatial statistics, mechanistic parameter calibration, correlation to outcomes) was implemented in Python using the following packages: numpy^58^, scipy^59^, scikit-learn^60^, shapely^61^, matplotlib^62^, and joblib^63^.

**Table 1:**
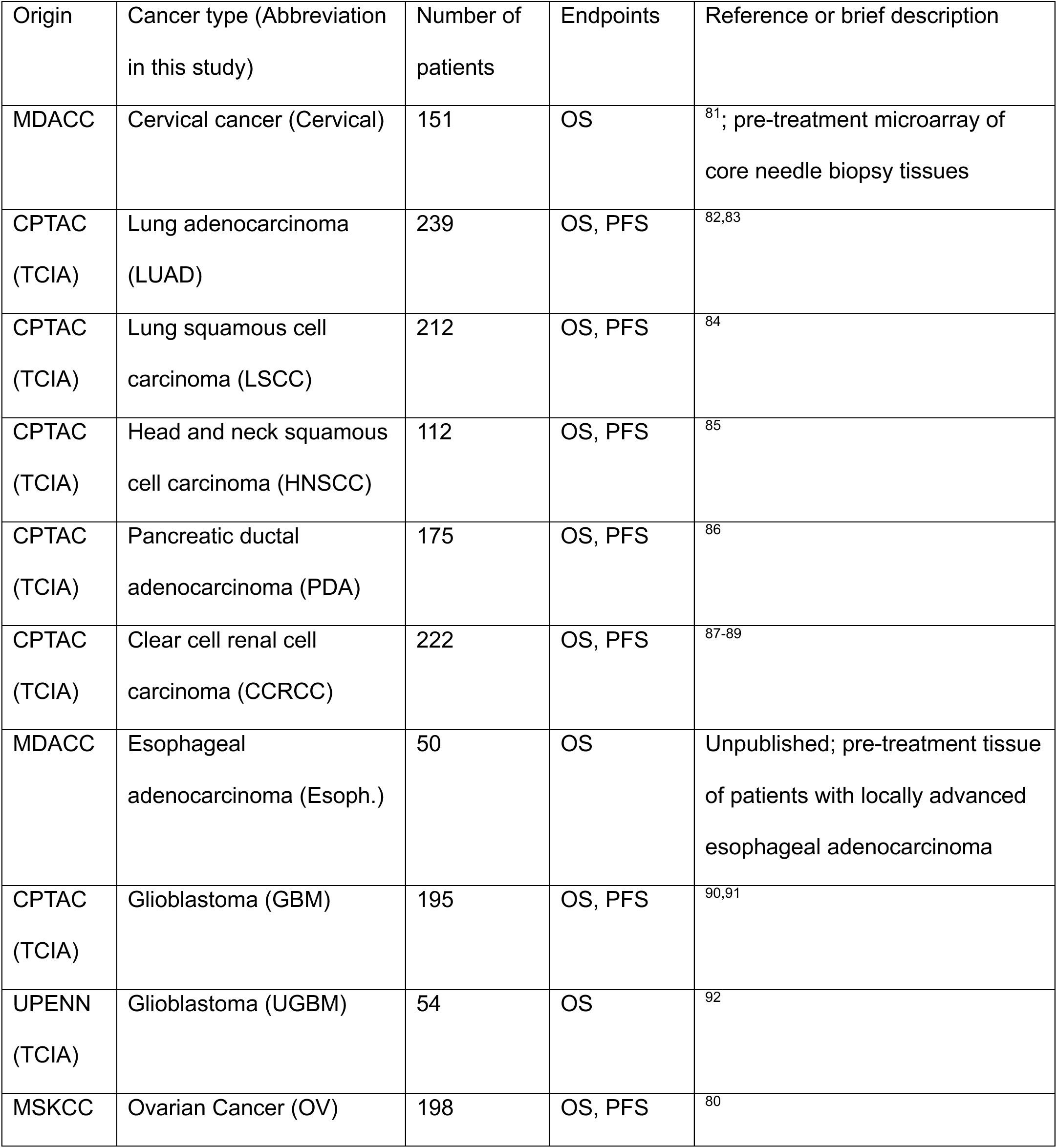
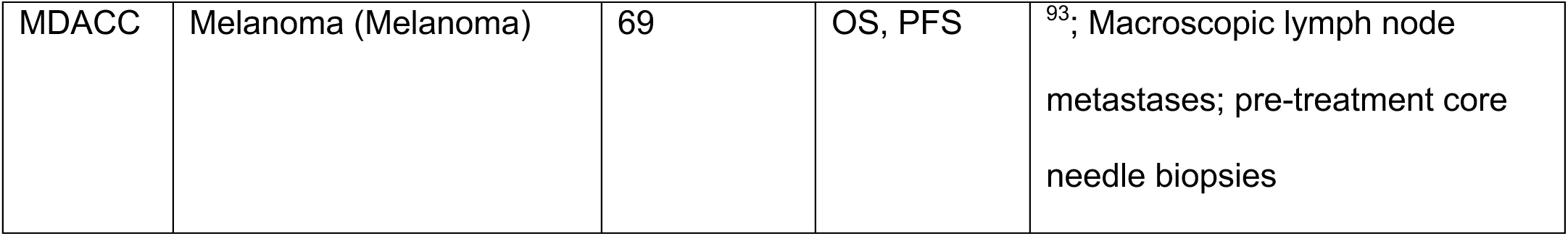
Dataset details.

| Origin | Cancer type (Abbreviation in this study) | Number of patients | Endpoints | Reference or brief description |
| --- | --- | --- | --- | --- |
| MDACC | Cervical cancer (Cervical) | 151 | OS | <sup>81</sup>; pre-treatment microarray of core needle biopsy tissues |
| CPTAC (TCIA) | Lung adenocarcinoma (LUAD) | 239 | OS, PFS | <sup>82,83</sup> |
| CPTAC (TCIA) | Lung squamous cell carcinoma (LSCC) | 212 | OS, PFS | <sup>84</sup> |
| CPTAC (TCIA) | Head and neck squamous cell carcinoma (HNSCC) | 112 | OS, PFS | <sup>85</sup> |
| CPTAC (TCIA) | Pancreatic ductal adenocarcinoma (PDA) | 175 | OS, PFS | <sup>86</sup> |
| CPTAC (TCIA) | Clear cell renal cell carcinoma (CCRCC) | 222 | OS, PFS | <sup>87-89</sup> |
| MDACC | Esophageal adenocarcinoma (Esoph.) | 50 | OS | Unpublished; pre-treatment tissue of patients with locally advanced esophageal adenocarcinoma |
| CPTAC (TCIA) | Glioblastoma (GBM) | 195 | OS, PFS | <sup>90,91</sup> |
| UPENN (TCIA) | Glioblastoma (UGBM) | 54 | OS | <sup>92</sup> |
| MSKCC | Ovarian Cancer (OV) | 198 | OS, PFS | <sup>80</sup> |
| MDACC | Melanoma (Melanoma) | 69 | OS, PFS | <sup>93</sup>; Macroscopic lymph node<br>metastases; pre-treatment core<br>needle biopsies |

### Nucleus Detection

The cell nuclei were automatically segmented from the raw histopathology slides^64^. The point patterns of segmented cell nucleus centroids were exported in (*x_i_*, *y_i_*) coordinate format in the distance unit *μm* for further processing.

### Point Pattern Preprocessing

For numerical stability, we mapped the raw centroid coordinates (*x_i_*, *y_i_*) to the first quadrant by subtracting the respective axis minimums. To ensure computational feasibility and contextual correction of cell conglomerates for subsequent steps, we identified connective cell clusters as individual representative entities on the respective slides. This is particularly important for slides with multiple tissues, such as core needle biopsies. For this purpose, HDBSCAN clustering was applied on the point patterns and downsampled to a maximum of 500,000 points using random uniform subsampling without replacement. If there were 500,000 points or fewer on one slide, all points were preserved for clustering. We then determined the concave hull of each identified cluster as the cluster boundary. All original (pre-downsample) points from one slide that fall inside this cluster hull are collected as an independent ROI ℛ. For each ROI, we determined the inner reference point, defined as the point closest to the respective ROI’s centroid. To ensure computational feasibility, the 5,000 points closest to this inner reference point were selected as samples for further spatial statistics processing. To ensure the accuracy subsequent spatial statistics, ROIs with fewer than 500 points were omitted. The number of points in one ROI ℛ is denoted as *N*_ℛ_.

### Pair Correlation Function

The area *A*_ℛ_ of an ROI was defined as the corresponding concave hull polygon area. All pairwise distances of points P*_i_* within an ROI were calculated, and the maximum pairwise distance defined as *r_max_* = max P*_i_*. The bin edges 0, …, *r_max_* were defined with a fixed step size Δr = 1 *μm*, such that the total number of individual bins is ⌊*r_max_*⌋. The number of pairs within the circle around the ROI centroid with radius *r*, *φ*(*r*), was identified. The counts of unique unordered pairs *c_k_* in each annulus [*r_k_*, *r_k_*_+1_) (with width Δ*r*) were determined using KD-tree cumulative neighbor counts and

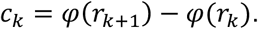

Since the ROI’s structure is highly dependent on the type of biopsy and pairwise distance calculation is only possible between points that have been observed on the biopsy, properties cannot be deduced from pairs that lie outside of the scanned tissue. This is particularly relevant for points closer to the boundary of the determined ROI, as it influences the proximity evaluations of cell centroid point patterns. To account for this, polygonal edge correction was performed. For each point *p_i_* we calculated the minimum Euclidean distance to the ROI boundary *d_i_*. For a given radius *r* the accessible fraction correction *e_i_*(*r*) is determined by

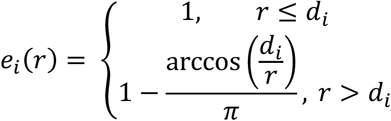

The number of counts of pairs 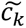 corrected for boundary truncation is defined by

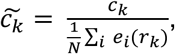

using the point density *ρ* = *N*_ℛ_⁄*A*_ℛ_, the corrected number of pairs was now used to determine the pair correlation function *g_RDE_*(*r*) for each cluster as

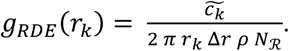

Gaussian smoothing was applied for the pair correlation function (over three distance bins) and omitted radii smaller than 10 µ*m* to mask close-proximity cell-cell interactions and focus on larger-scale tissue effects. The rescaled pair correlation function *g*(*r*) was calculated on the relative radii *r_k_*⁄*r_max_* to ensure comparability between different biopsy modalities.

### Transformation of Pair Correlation Function to Power Spectral Density

To transform the 2PCF *ξ*(*r*) = *g*(*r*) − 1 to the PSD *P*(*k*), and matching the already used assumptions of isotropy, we employed the radial symmetric Hankel transformation with Bessel’s *J*_0_. This results in the closed form expression for a frequency *k* in two dimensions,

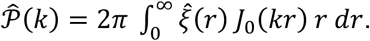

For the inverse transformation used to determine the 2PCF from the PSD in subsequent steps, we used the analytical expression

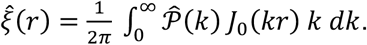

The Hankel pair integrals are evaluated with trapezoidal quadrature on a logarithmic-spaced *k* grid.

### Joint Calibration of Mechanistic Parameters

We identified the parameter pair (*γ*, *D*) by nonlinear least squares fitting on the concatenated observation vector with *n* bins for the 2PCF and *m* bins for the PSD:

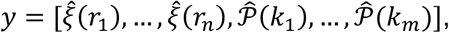

which enabled parallel fitting of the observed characterizations in both distance and frequency space to determine the dynamical model parameters. The parameter-dependent corresponding concatenated model formulation using the analytical derivations from above on the normalized time scale reads

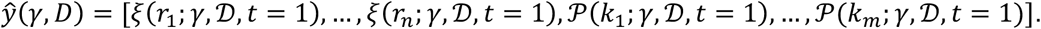

Positivity of the two estimated parameters was enforced by optimizing for ln *γ* and ln D. The corresponding standard errors were derived from the asymptotic covariance of the least squares estimator.

### Goodness of Fit

For each ROI, we determined the goodness-of-fit coefficient of determination *R*^2^ separately for the 2PCF and PSD components:

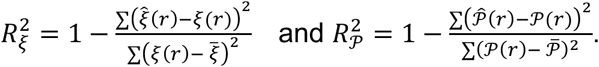

For reference, we also calculated the unweighted average of these two metrics as representative measure 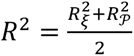 for every single fit. Corresponding normalized root mean squared errors (NRMSEs) were calculated separately for 2PCF and PSD as the root mean squared deviation between data-derived and fitted curves, normalized by the dynamic range of the respective data-derived curves:

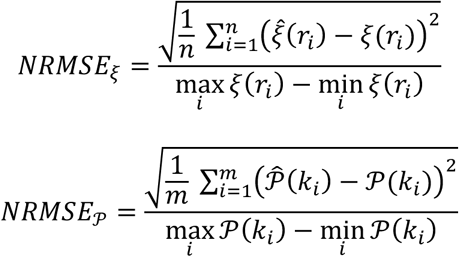

These metrics were reported, using mean and standard deviation, for each ROI across cohorts.

We also compared this joint fitting approach with fitting on only the PSD^34^. All goodness-of-fit metrics were recalculated analogously and their values compared with the joint fitting approach. Differences in performance were assessed using the two-sided paired Wilcoxon signed-rank test (i.e., the explicit null hypothesis was whether the median of differences in performance was equal to zero).

### Profile Likelihood Analysis for Per-ROI Parameter Identifiability

One-dimensional profile likelihoods were used for the fitted parameters *γ* and D to quantify parameter identifiability for each individual ROI. The joint least-squares estimate is given by minimizing the combined residual:

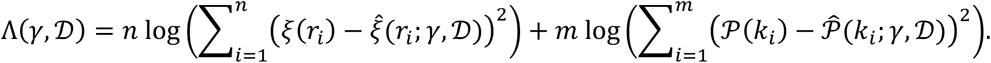

The corresponding scaled likelihood ratio for a candidate parameter pair (*γ*, D) can be written as

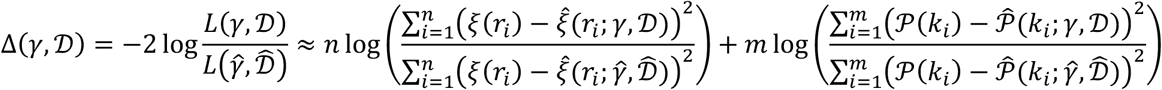

under the working assumption of independent Gaussian errors with unknown but constant variances. For each parameter θ ∈ {*γ*, D}, the one-dimensional profile likelihood was constructed after log-scaled parametrization over the domain of definition. This procedure was performed for the joint fit (2PCF and PSD) as well as the PSD-only fit (i.e., omitting the *ξ*-terms) for comparison. Approximate 95% confidence intervals (CIs) for *γ* and D were obtained from the profile likelihood through identifying the grid points (*γ*_j_, D_j_) on the discretized domain of definition that had a 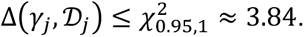 The boundaries of this set of grid points were defined as the CI of reference, and the relative fraction of the greater to the smaller boundary was subtracted from 1 to derive the relative CI width, which was used to determine whether joint fits provided narrower CIs than PSD-only fits. To visualize and provide a descriptive summary of the variability of profile likelihoods across ROIs, aggregations of the per-ROI profile likelihoods were constructed.

### Use of Derived Parameters for Microscopic Simulations

To visualize how different combinations of the two calibrated kinetic parameters *γ* and D manifest in microscopic tissue architectures, synthetic point patterns were generated. In a Thomas point process, parent points are distributed according to a homogeneous Poisson process with intensity *κ*, and each parent independently generates a Poisson-distributed number of offspring points with mean *μ*. The offspring points of a given parent point are placed around the parent location according to an isotropic bivariate normal distribution with standard deviation *σ*. The Thomas process has constant intensity *λ* = *κ μ* and a closed-form pair correlation function

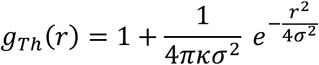

for inter-point distance *r* in two dimensions^65–67^. To ensure that the forward simulations are consistent with the reaction-diffusion model used for parameter calibration, we linked the Thomas process parameters *κ* and *σ* directly to the inferred kinetic parameters *γ* and D, for which we utilized the analytical 2PCF of the reaction-diffusion equation *g*(*r*)*_RDE_*.

By equating the width and amplitude of this pair correlation function with the known Thomas pair correlation function, we obtain 4*σ*^2^ = 8D*t*, i.e., 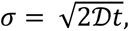 and 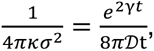 i.e., *κ* = *e*^−2^*^γt^*. At the above introduced normalized time scale, this yields the mapping 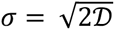 and *κ* = *e*^−2^*^γ^*. This identification is derived directly from the two closed-form expressions and makes the Thomas process 2PCF coincide with the reaction-diffusion 2PCF at the normalized time scale.

For each selected pair of parameters *γ* and D, we fixed a quadratic domain as reference to the observed histology patches. Given *κ* from the mapping above, the mean offspring number was defined as *μ* = *λ*/*κ* for *λ* fixed and a Thomas process was simulated. Parent points were drawn from a homogeneous Poisson process with intensity *κ*, and offspring points were generated for each parent and displaced by independent bivariate normal vectors with standard deviation *σ* in both spatial directions. The resulting point patterns represent synthetic nucleus centroids. From these simulated point patterns, the pair correlation function was determined and the estimates compared with the analytical Thomas pair correlation function as well as the analytical reaction-diffusion pair correlation function for verification.

### Use of Derived Parameters for Macroscopic Simulations

The calibrated parameters *γ* and *D* were utilized to simulate the reaction-diffusion equation on a Cartesian grid to obtain macroscopic *in silico* tumor-cell densities. The spatial Laplacian was discretized on a square domain with Dirichlet boundary conditions, and time integration was performed with operator splitting. A semi-implicit reaction step according to *u* ↦ *ue^γ^* ^Δ*t*^ (or explicit Euler for *u_t_* = *γu*) was followed by an implicit diffusion step *u* ↦ (*I* − D Δ*t* Δ)^−1^ *u*, where the linear system was solved via sparse LU factorization and reused for all simulations at fixed D. This produced smooth density profiles *u*(*x*, *t*) and used the same kinetic parameters as those used for the Thomas point process simulations. This enabled microscopic (point-pattern) and macroscopic (density) visualizations in the *γ*-D space, as well as in the derived parameter-combination spaces.

### Construction of the Pan-Cancer Parameter Map

To visualize the parameter distributions across cohorts, a pan-cancer map was constructed in transformed parameter space. The individual estimates of *γ* and D were used for each patient. Before pooling across datasets, the parameters were standardized at the global (pan-cancer) level by computing z-scores:

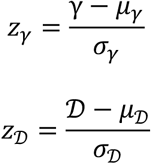

where *μ_γ_*, *σ_γ_*, *μ*_D_ and *σ*_D_ denote the mean and standard deviation of *γ* and D calculated over all patients, respectively. An orthogonal system of coordinates (*x*, *y*) was obtained by defining the linear combinations *x* = *z_γ_* + *z*_D_ and *y* = *z*_D_ − *z_γ_*. In this representation, *x* increases when both *γ* and D are high relative to the global mean, whereas *y* captures whether a tumor is relatively more diffusion-dominated (high D, low *γ*) or proliferation-dominated (high *γ*, low D). For each cohort, the centroid in this coordinate system was computed as the arithmetic mean of all patients from the respective dataset. This enabled the comparison of the cohort-specific parameter distribution centroids between cohorts. Analogous transformations were performed within the respective cohort’s parameter values to identify intra-cohort parameter heterogeneities.

### Extension of the Mechanistic Phenotypes: Dispersion Length and Front Velocity

From the two identified kinetic parameters *γ* and D that can be interpreted as phenotypic characterizations, we further derived two composite metrics with direct mechanistic interpretations: dispersion length 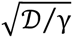 and front velocity 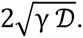. These established characteristics of reaction-diffusion equations were used to characterize the mechanistic phenotypes because they define the relationship between *γ* and D.

### Survival Analysis of Phenotypes Encoded in Parameter Space Tiles

For each cohort and endpoint, the cohort-specific two-dimensional dispersion-length and front-velocity parameter space was subdivided into a regular 3×3 grid. The boundaries of the resulting nine tiles were defined as the tertiles of each parameter’s distribution, such that each tile corresponded to low, intermediate, or high values of each parameter on the cohort-specific scale. For each tile, we recorded the number of patients, the number of events, the event rate (the proportion of patients with an event, ignoring censoring), and the Kaplan-Meier estimate of the median time to event. To formally test whether survival differed across phenotypes in this grid, a global log-rank test compared the Kaplan-Meier survival curves for all tiles per cohort and endpoint. Tiles with fewer than eight patients and three events were excluded from the respective test to avoid unstable estimates. The resulting p-values should be interpreted as evidence of cohort-level variation in survival across the phenotypic space. The tile-specific event rates and median survival times were visualized as heatmaps over the parameter grid. Per cohort and endpoint, a single global log-rank test was performed; no multiplicity adjustment was applied across cohorts or endpoints, as each cohort is reported and interpreted as an independent validation^68^.

### Univariate Kaplan-Meier Analysis Stratifying for Identified Parameters

We further evaluated associations between the respective identified patient-specific parameter values and outcome. For each cohort, we performed tertile splits to compare overall and progression-free survival using log-rank tests for group comparisons to identify potentially relevant differences. In this exploratory analysis, p-values less than 0.05 were considered indicative of an association between the respective parameter and outcome.

### Orthogonality of Information to Anatomical Stage in CPTAC Cohorts and Added Clinical Value in Glioblastoma

CPH models were fit for PFS in the GBM cohort (events-per-variable-ratio 13.0). The baseline CPH model used age (continuous and z-scored) and sex (binary). The HDTD-augmented CPH model adds the log-transformed HDTD pair (log *γ*, log D) to the baseline model. Ridge penalization (*λ* = 1.0) was applied when events per variable dropped below five^69^. Reporting of the multivariable Cox development and performance metrics follow the TRIPOD+AI statement for prediction-model studies^70^. Nested-model likelihood-ratio tests quantified added information. Harrell’s c-index^71^ and its bootstrap 95% CI (1000 patient-level resamples of patients with the fitted model held fixed) quantified discrimination. Continuous net reclassification improvement and integrated discrimination improvement at 12 months were computed with bootstrap 95% percentile CI derived from 1000 patient-level resamples and both nested models refit at each resample^72^. Calibration at 12 months was assessed by binning patients into five equal-sized bins of predicted event probability and comparing the mean predicted event rate per bin against the Kaplan-Meier observed event rate. Proportional-hazards assumptions per covariate were examined via the *χ*-squared statistic of the rescaled Schoenfeld residuals^73,74^. Orthogonality of HDTD parameters to anatomical Stage was assessed in the five CPTAC cohorts with standardized TNM staging using partial Spearman rank correlation residualized on age and sex, and pooled across cohorts via inverse-variance weighted Fisher-z transformation. *I*^2^ was used to quantify between-cohort heterogeneity^75^. Glioblastoma was excluded in the orthogonality analysis since standardized TNM Stage is not available.

### Histology-Calibrated *in silico* Radiotherapy Counterfactuals in Glioblastoma

To showcase the potential applications of histology-derived kinetic parameters in treatment-specific forward simulations, mechanistic reaction-diffusion equation simulations for representative phenotypes in the glioblastoma cohort were performed. Four glioblastoma parameter combinations were selected spanning high versus low effective proliferation rates *γ* and diffusion coefficients D, derived from the cohort-specific distribution of calibrated parameters. For each pair (*γ*,D), the radially symmetric reaction-diffusion equation 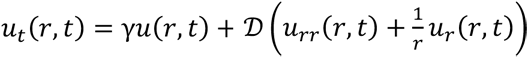 was solved in polar coordinates and numerically on a circular domain using finite-difference discretization in space and explicit time steps consistent with the histology-based calibration. HDTD calibrates dynamics on a normalized scale; thus, absolute calendar-time mapping was a modeling choice and was put into perspective for known orders of magnitude^76^. Absolute times should be interpreted as illustrative. Tumors were initialized as a small central core and grown until the diameter of the isocline *u* ≥ *u_dia_*_g_ reached a predefined macroscopic “diagnosis” diameter of 5 *mm* above an assumed tumor density detection threshold of 20%, at which time point the state was defined as baseline for treatment.

From this diagnosis state, an idealized planning target volume (PTV) was applied, covering the entire domain of definition for *in silico* radiation. Radiotherapy was modeled using a linear-quadratic (LQ) formalism applied to the PTV. The standard of care was simulated as 2 Gy once daily to 60 Gy in 30 weekday fractions (six weeks of treatment). The LQ parameters were linked to the calibrated kinetics^15^ by setting *α* = *k_α_ γ* with *k_α_* = 1 and a fixed α/β = 10 Gy. The survival fraction was set to 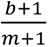 for each fraction dose *d*, which was multiplicatively applied to the cell density within the PTV at the corresponding time steps. Between fractions, spatial diffusion and proliferation were simulated according to the same (*γ*, D) as in the untreated system. For each forward simulation, we recorded total tumor burden, defined as the spatial integral of *u*(*r*, *t*) over the domain. The densities directly after the last fraction were compared to assess treatment efficacy. To identify schedules that map the low proliferation phenotypes onto the post treatment state of the fast proliferating one under standard of care, candidate schedules with fraction sizes between 1.8 and 3.4 Gy were considered with biologically effective doses between 50 and 100 Gy. They were forward simulated for the low proliferating phenotypes and ranked by the relative L2 distance.

### Counterfactual Schedule Allocation in an *In Silico* Trial in the Glioblastoma Cohort

Counterfactual event times were calculated by shifting each observed time by the simulated schedule contrast and re-censoring^35^, such that the observed cohort serves as the reference arm. A counterfactual time of event can become non-positive when slow proliferating tumors are given a much shorter treatment course. This yields unfeasible values of event times, and to prevent these situations, the treatment deescalations can be given only to faster proliferating tumors. For the slower proliferating ones, an escalation only adds time, and the problem does not occur. The patients are taken to have received the standard of care for newly diagnosed glioblastoma over the accrual period, 60 Gy in 30 fractions of 2 Gy with concomitant and adjuvant temozolomide^77^. The different treatment strategies were compared by restricted mean survival time truncated at 12 and 24 months, and by median survival. No log-rank test was performed and no hazard ratio reported since the compared arms are counterfactuals on the identical patients rather than independent groups. The effect size is bounded by a bootstrap over patients as sampling unit, and the hypothesis that the proliferation parameter carries no allocation information is tested by keeping the treatment schedules of the arm with stratification and permuting which patients receive which of the two courses, each patient maintaining their own modeled delay under the course they are given, so that both arms remain feasible schedules. The cohort was split at the median and every interval rests on *m* = 2,000 resamples, with the permutation p value defined as 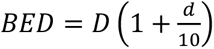 with *b* the number of random allocations containing the observed difference. The split threshold, the arm assignment and the best single comparator are all estimated from the identical patients, so the bootstrap re-runs the entire procedure rather than just resampling the two final curves. Schedule contrasts are not evaluated with the partial differential equation used for the phenotype simulations above but with an analytical closed form. The calibration normalizes the time scale such that the calibrated rate *γ* absorbs the unknown interval between tumor onset and the time of biopsy. Reformulating this interval as a power of the rate and fixing the median of the cohort leaves the spread free and returns the absolute rate

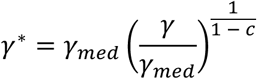

with an assumed compression exponent c=0.98. Radiosensitivity is coupled to this via

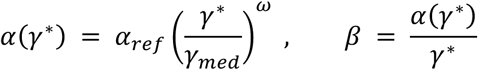

with a coupling exponent *ω* = 0.25 and a reference radiosensitivity of 0.289 per Gy, the value implied by a surviving fraction of 0.50 at 2 Gy for an *α*/*β* ratio of 10 Gy. The delay that a schedule k confers relative to the reference course is then

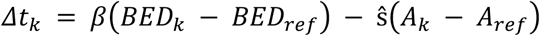

where 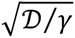 is the biologically effective dose of a total dose *D* given in fractions of *d* Gy. The number of calendar days of the course (weekends left untreated) is denoted by *A*. The patient-specific detection threshold and reference course both cancel from this, which is why only differences between schedules are reported. Across the six simulated uniform schedules (75 Gy in 30, 70 Gy in 35, 60 Gy in 30, 50 Gy in 20, 40 Gy in 15 and 30 Gy in 5 fractions) the *BED* is 93.8, 84.0, 72.0, 62.5, 50.7 and 48.0 Gy and *A* is 40, 47, 40, 26, 19 and 5 days, respectively. Per day of protraction a delay of ŝ = 2.87 days is assumed. The coefficient for tumor regrowth is the reciprocal of the dose that offsets the repopulation occurring in one day, reported for glioblastoma as 0.30 Gy per day (0.22-0.39) at 2 Gy per fraction. Expressed per Gy of biologically effective dose the coefficient is identified to lie in an interval from 2.05 to 3.64 days per Gy^78^. Its midpoint, 2.84 days per Gy, multiplied by the published time factor of 0.66 Gy per treatment day for accelerated repopulation^79^ gives 1.87 days per treatment day, and an additional day per treatment day is added for the treatment day itself, on which the model produces no regrowth. Since every reported quantity is a difference between two schedules simulated on the identical patient, the declared reference course cancels from the delay contrasts. It does not cancel from the reported restricted mean difference, as re-censoring and truncation operate on the shifted times. Declaring every single of the candidate schedules in turn as the course the cohort received left the allocation identical and moved only the gain against the standard of care.

### Statistical Analysis

All analyses were performed in Python as described. No statistical method was used to predetermine sample size, since sample sizes corresponded to the number of patients and slides available in each cohort (see Table 1). ROI-level parameter estimates were patient-specifically aggregated into single-patient-level metrics using the ROI-level medians. The parameters were estimated per ROI by joint nonlinear least-squares fitting of the two-point correlation function and the power spectral density to their analytically derived model counterparts on a normalized time scale. Goodness of fit was quantified using the coefficient of determination and normalized root mean squared error for 2PCF and PSD separately and as an unweighted mean of both. Practical parameter identifiability was assessed by one-dimensional profile likelihoods. The performance of joint versus PSD-only fitting was compared using the two-sided paired Wilcoxon signed-rank tests. Time to event outcomes were analyzed using Kaplan-Meier estimates and compared between groups using two-sided log-rank tests. This does not apply for the counterfactual schedule comparison, where arms are not independent and a permutation test on the allocation was used instead. For the tiled phenotype analyses, cohort-specific subgroups were defined by tertiles of the respective parameter dimensions, and tiles with fewer than eight patients and fewer than three observed events were excluded. In exploratory univariate analyses, tertile splits were used to conduct global log-rank tests, and a significance level below 0.05 was interpreted as indicative of association.

## Supporting information

Supplementary Figures (zip archive)

## Data availability

This study is a secondary analysis of already existing, de-identified human data. The whole-slide images and the clinical records were collected, consented and released by the Clinical Proteomic Tumor Analysis Consortium and the associated repositories under their own approvals, and are publicly available as cited in the Data availability statement. No new human data were collected. Unless otherwise stated, the data sets analyzed in this study are published, referenced, and publicly available online through the referenced locations. Some data used in this publication were generated by the National Cancer Institute Clinical Proteomic Tumor Analysis Consortium (CPTAC). The CPTAC whole-slide imaging data analyzed in this study are publicly available through The Cancer Imaging Archive (TCIA):

CCRCC (https://doi.org/10.7937/k9/tcia.2018.oblamn27),

GBM (https://doi.org/10.7937/K9/TCIA.2018.3RJE41Q1),

HNSCC (https://doi.org/10.7937/K9/TCIA.2018.UW45NH81),

LSCC (https://doi.org/10.7937/K9/TCIA.2018.6EMUB5L2),

LUAD (https://doi.org/10.7937/K9/TCIA.2018.PAT12TBS),

and PDA (https://doi.org/10.7937/K9/TCIA.2018.SC20FO18).

UPenn-GBM data are available through TCIA (https://doi.org/10.7937/TCIA.709X-DN49). Ovarian cancer data associated with Boehm et al.^80^ are available through the original publication. The de-identified institutional clinical data sets are available from the corresponding author Heiko Enderling and the respective data owners upon reasonable request with completion of data-sharing agreements, subject to institutional and regulatory policies.

## Code Availability

Code implementing the presented method can be found in the GitHub repository: https://github.com/psch-mdacc/HDTD.

## Acknowledgements

The authors acknowledge the High-Performance Computing for Research facility at The University of Texas MD Anderson Cancer Center for providing computational resources that have contributed to the research results reported in this paper. Pirmin Schlicke would like to thank Shambhavi Kurup for helpful feedback.

Pirmin Schlicke was supported by a grant from the Radiation Oncology Strategic Initiatives (ROSI) of The University of Texas MD Anderson Cancer Center, Division of Radiation Oncology and in part funded within the APART-USA program of the OeAW. Caner Ercan was supported by the Swiss National Science Foundation (SNSF; P500PM_214162). This work was supported in part by the Jayne Koskinas Ted Giovanis Foundation for Health and Policy, a Maryland private foundation dedicated to effecting change in health care for the public good, to Heiko Enderling. The opinions, findings, and conclusions or recommendations expressed in this material are those of the authors and do not necessarily represent the official views of the Jayne Koskinas Ted Giovanis Foundation for Health and Policy, its directors, officers, or staff. Ashlee N. Seldomridge has support provided by the National Institutes of Health (United States NIH T32 CA009599 and NIH P30-CA016672 MD Anderson Cancer Center Support Grant). Ashley M. Holder was supported by the American College of Surgeons Clowes Award and the Department of Defense CDMRP Melanoma Academy Scholar Program. Francisco Vega was supported by funds from the T/NK Cell Malignancy Moon Shot Program at The University of Texas MD Anderson Cancer Center, P01CA272295-01 (NCI), U54CA302435 (NCI), and the Leukemia & Lymphoma Society (SCOR 7038-25). The funders had no role in study design, data collection, analysis or preparation of the manuscript.

## Author contributions

Conceptualization: PS, HE

Data acquisition & curation: PS, ANS, SA, AM, TPS, PF, TCN, AK, LC, AMH, RW, SHL, CATC, FV

Methodology: PS, CE, SR, MUZ, VAH, SP, CK, HE

Project administration: PS, HE Resources: CATC, FV, YY, HE

Validation: PS, CE, SA, ANS, AM, TPS, PF, NPO, FV

Writing - original draft: PS

Writing - review & editing: all authors

