## Supplementary figures and images for "Effective tumor kinetics inferred from single routine H&E biopsies enable counterfactual virtual radiotherapy trials"

### Supplementary_Figure_1.png

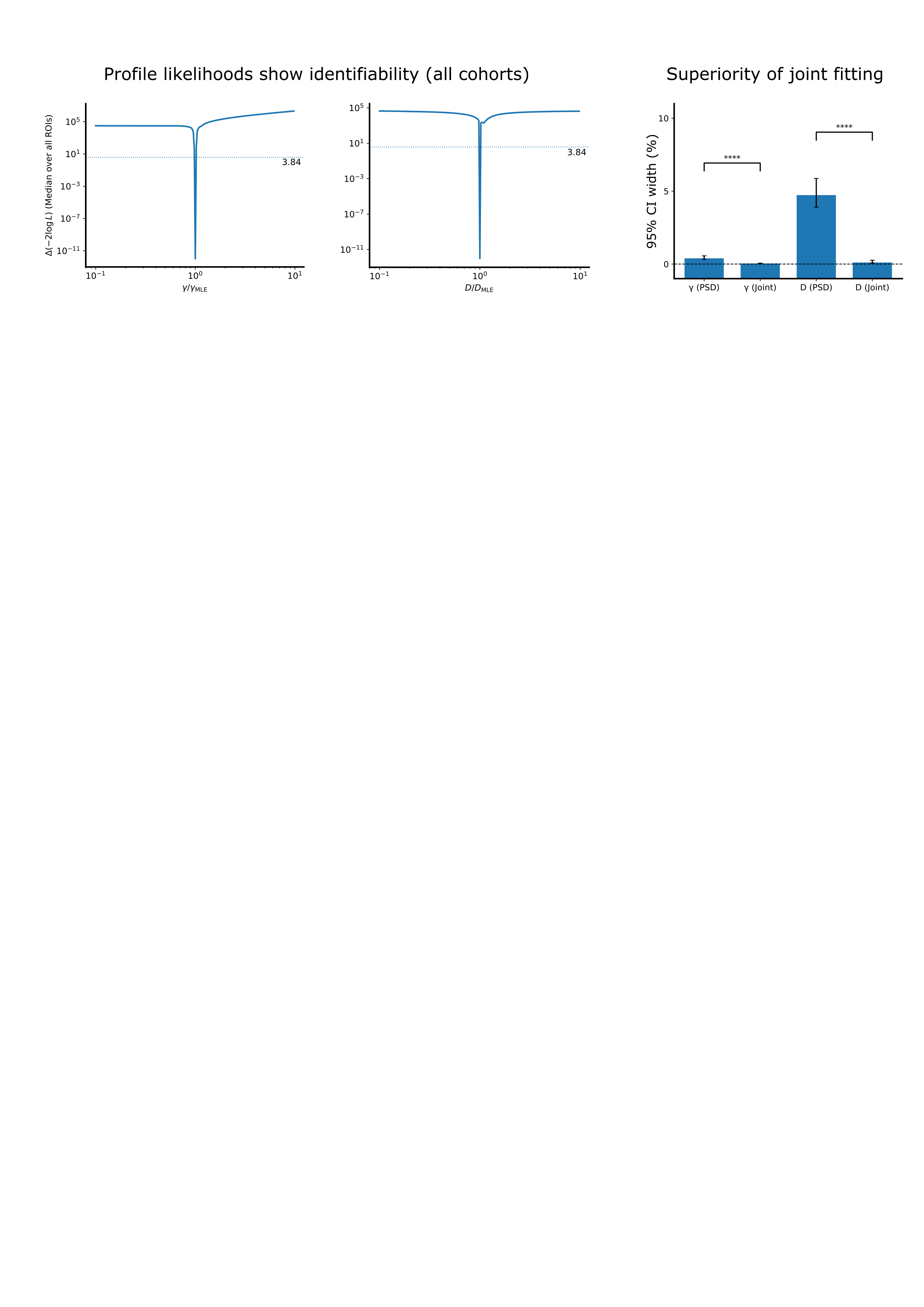

### Supplementary_Figure_2.png

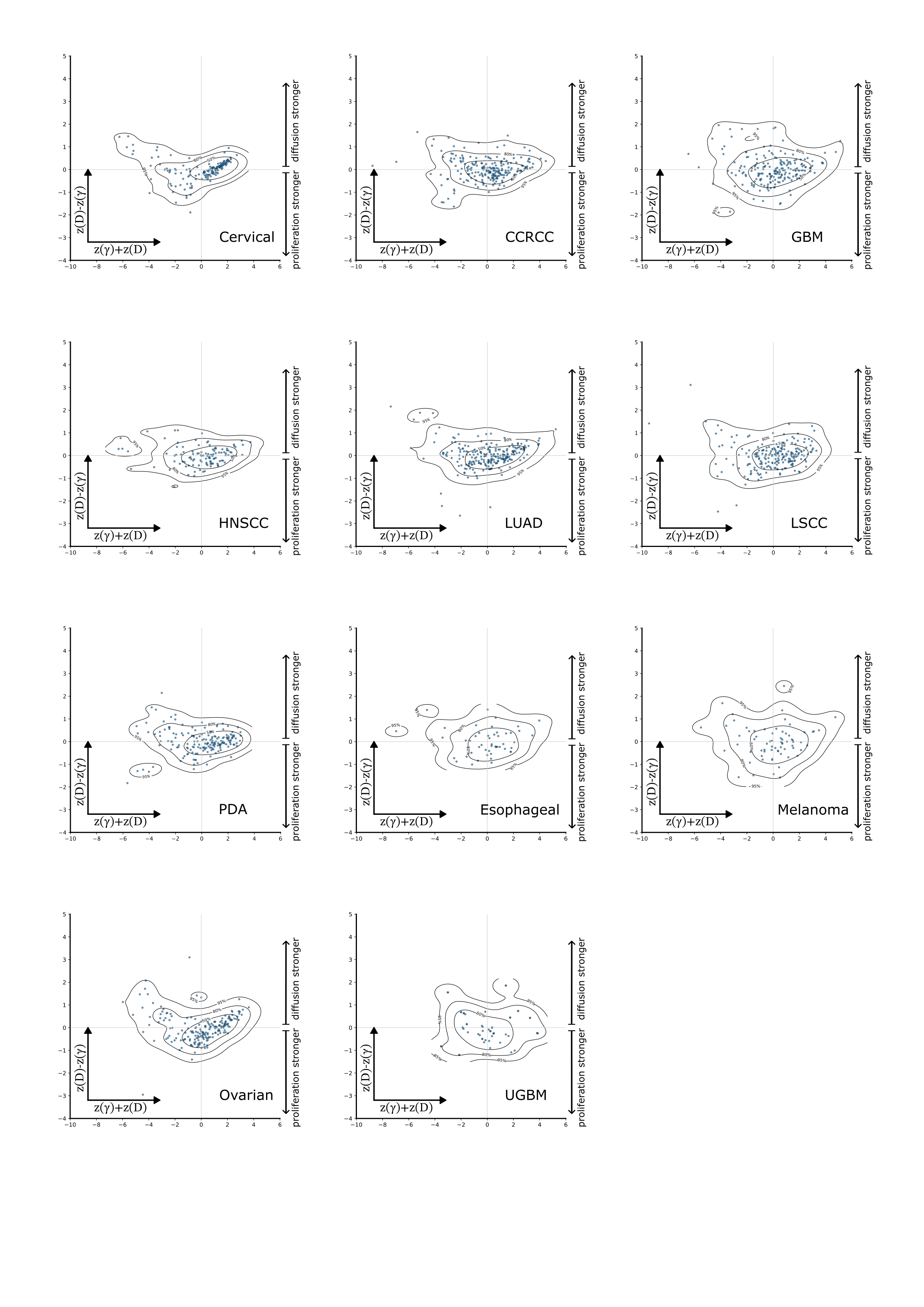

### Supplementary_Figure_3.png

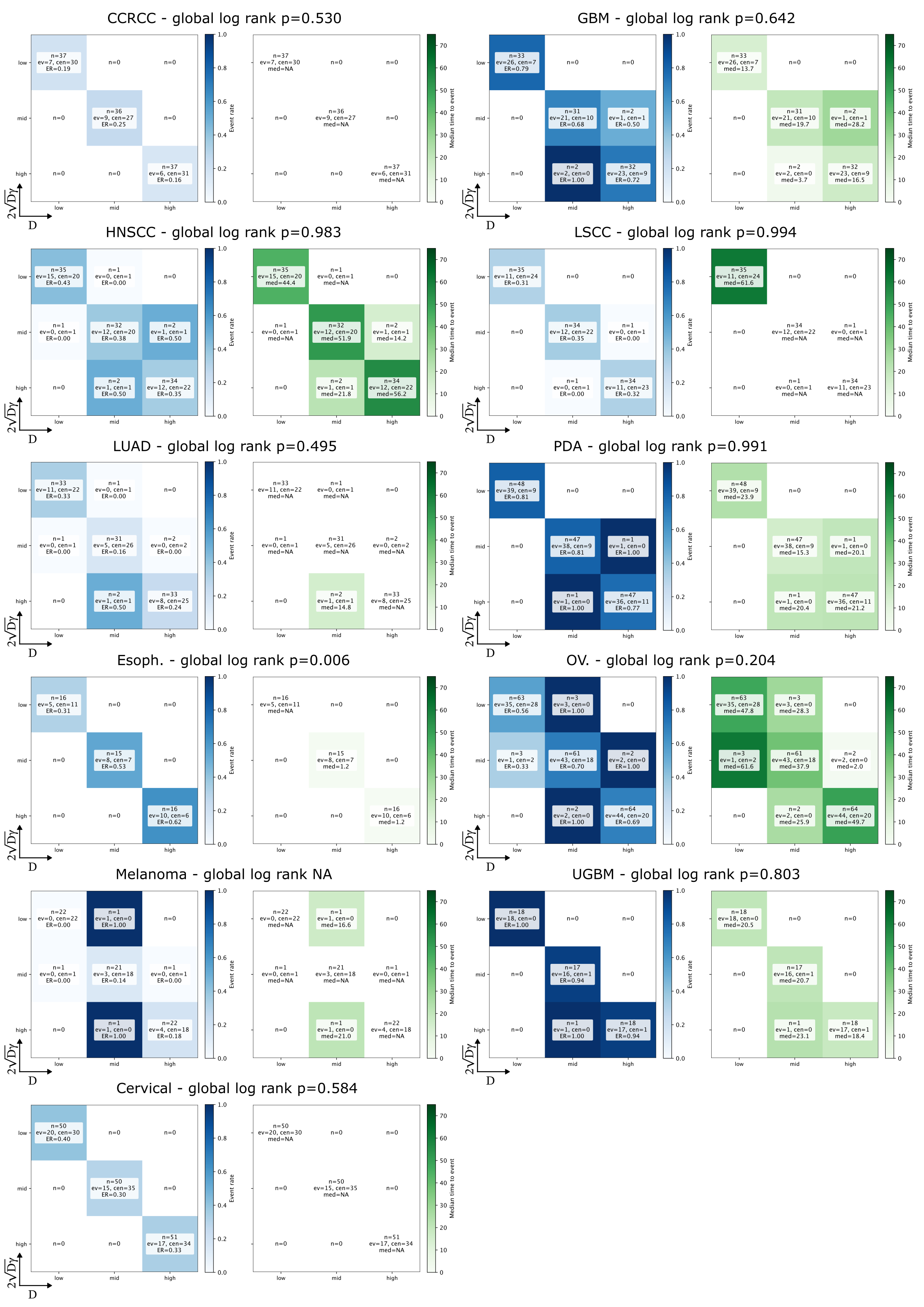

### Supplementary_Figure_4.png

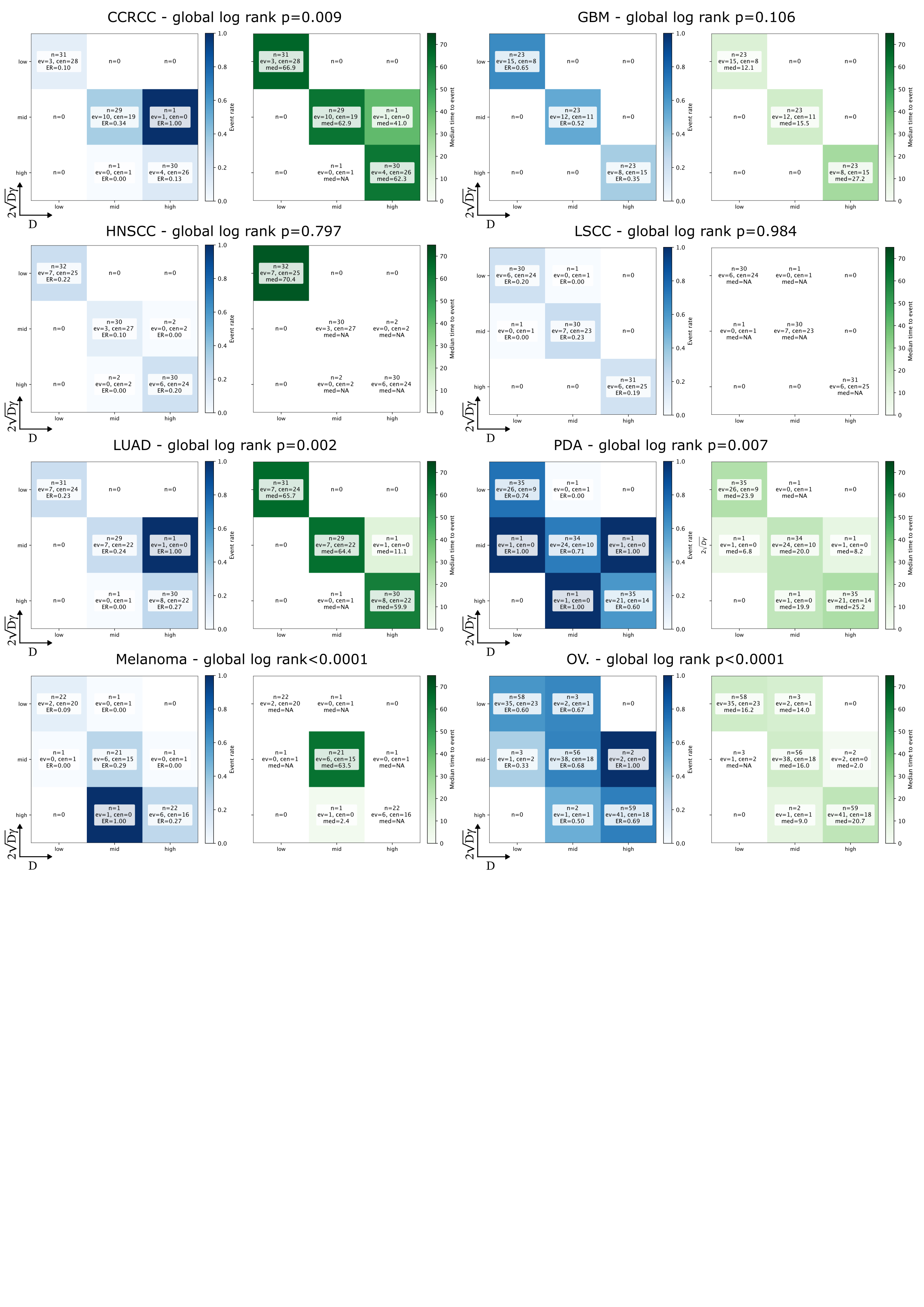

### Supplementary_Figure_5.png

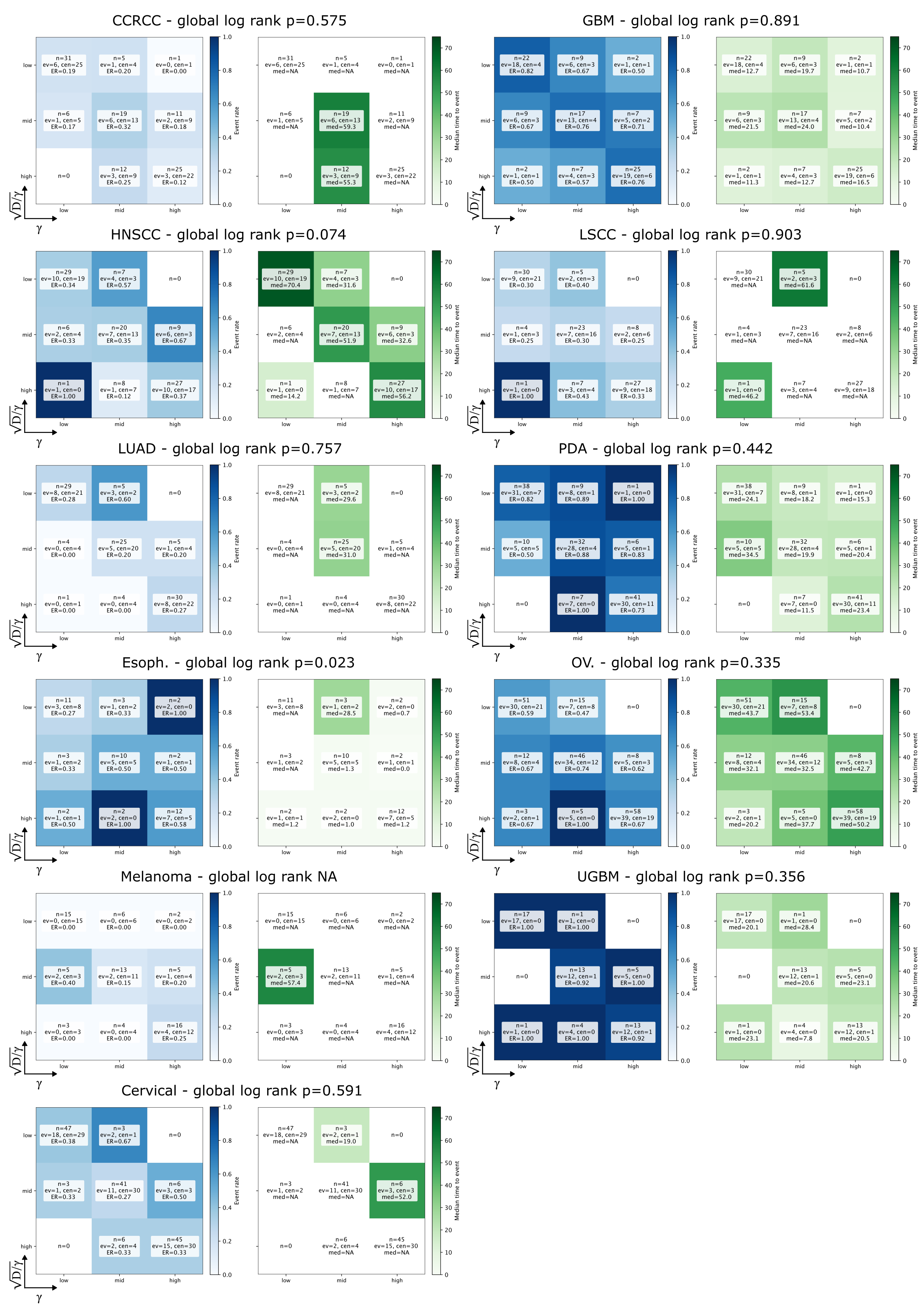

### Supplementary_Figure_6.png

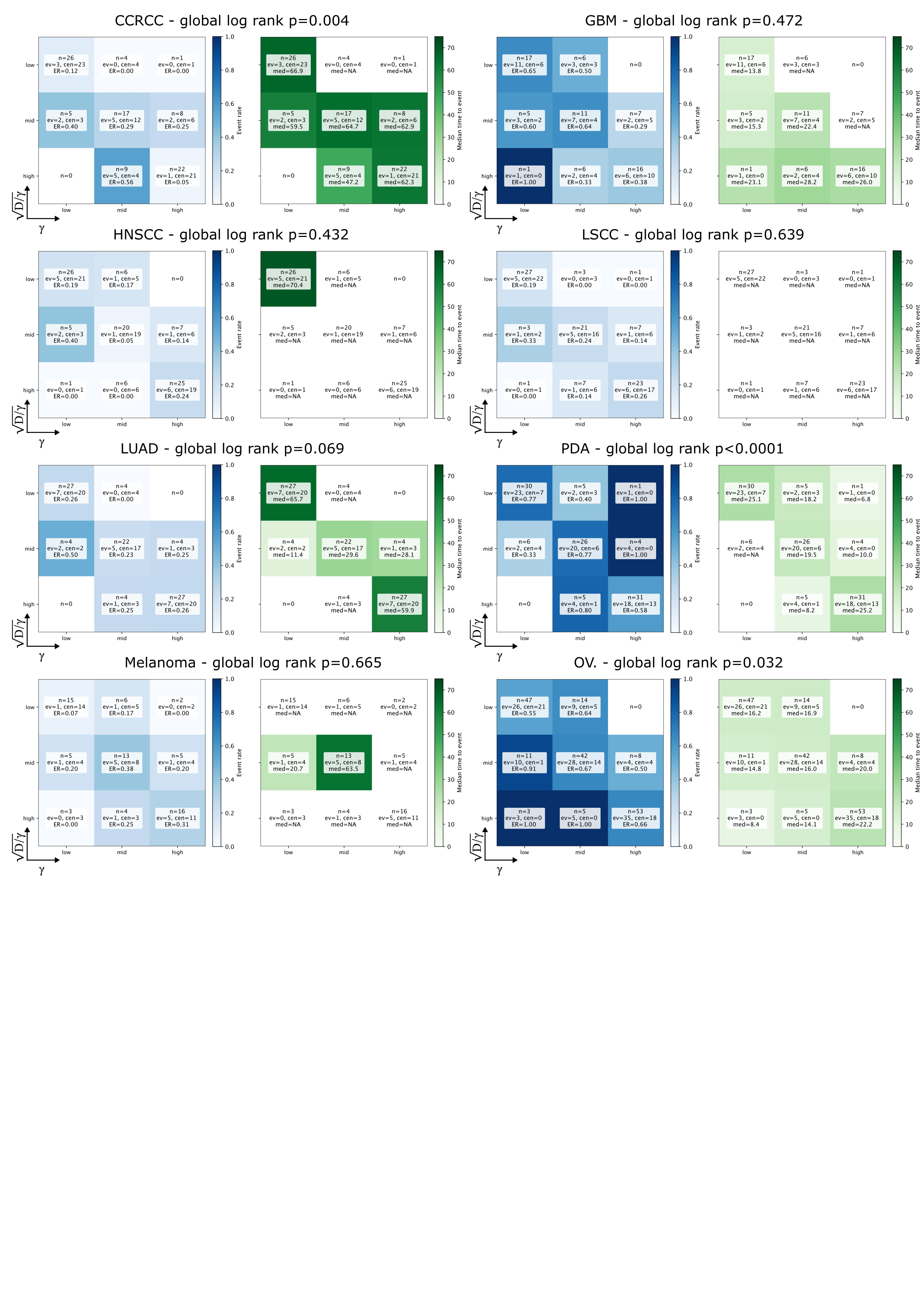

### Supplementary_Figure_7.png

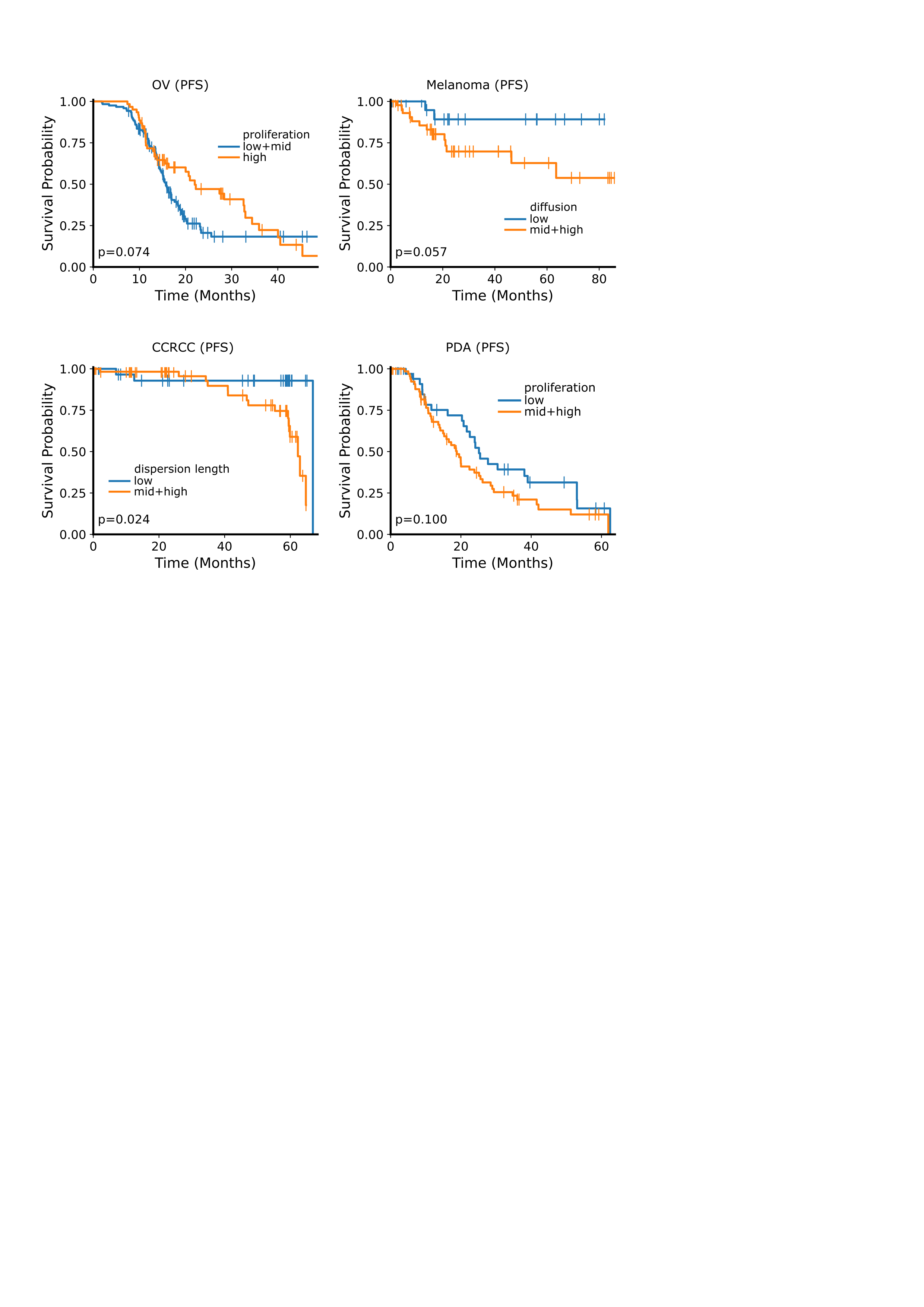
